# Host Genetic Regulation of NLRP3 Inflammasome Cytokines Reveals Immune and Vascular Pathways in HIV

**DOI:** 10.64898/2026.06.08.26355202

**Authors:** Ryan Chung, Namratha Shivani Chalasani, Alton S. Barbehenn, Erik Lundgren, Sonia Savur, Sayane Shome, Caroline H. Sheikhzadeh, Sannidhi Sarvadhavabhatla, Maria Sophia Donaire, Vivian Pae, Xiuping Chu, Daniel Winder, Colin T. Maguire, Senay Topal, Anuradha Ganesan, Joseph M. Yabes, Derek T. Larson, Tahaniyat Lalani, Evan C. Ewers, Rhonda E. Colombo, Eli Dugan, Ujjwal Rathore, Alexander Marson, Brian K. Agan, Jeffrey A. Tomalka, Rafick P. Sekaly, Nilah M. Ioannidis, Sulggi A. Lee

## Abstract

People with HIV exhibit elevated inflammation and cardiovascular risk despite antiretroviral therapy. To define the genetic architecture of inflammasome-associated inflammation, we performed whole-genome sequencing and quantified plasma IL-6, IL-1β, and IL-18 in 1,000 ART-suppressed PWH from the U.S. Military HIV Natural History Study. Genome-wide analyses identified 14 loci implicating antiviral defense (*DDX17, DDX41, EEA1, BCL11A*), lipid metabolism (*ABCA1, ABCA12, ABCC1, AGMO*), and vascular remodeling (*KLHL29, RNF213, ETV1*). Transcriptome-wide analyses across cardiovascular and immune tissues identified regulatory programs linking interferon signaling, immune activation, and vascular biology to circulating cytokine levels. Mendelian randomization analyses supported causal relationships between inflammasome-associated cytokines and vascular events. Functional integration with genome-wide CRISPR perturbation datasets in primary CD4⁺ T cells linked cytokine-associated loci to HIV antiviral pathways and cytokine regulatory networks. External validation in cohorts without HIV demonstrated pathway-level convergence despite limited variant-level overlap. These findings define genetic mechanisms linking inflammasome signaling, antiviral defense, and cardiovascular risk.

## INTRODUCTION

Systemic inflammation and immune activation are increasingly recognized as key drivers of atherosclerotic cardiovascular disease (ASCVD), alongside traditional risk factors.^1,2^ ASCVD remains a leading cause of global morbidity and mortality,^3,4^ and the mechanisms linking chronic inflammation to vascular disease are incompletely understood.^5,6^ This gap is particularly evident in people with HIV (PWH), who, despite effective antiretroviral therapy (ART), exhibit persistent immune activation and elevated risks of cardiovascular morbidity and mortality compared to people without HIV (PWoH).^7–15^ HIV infection is also prothrombotic state and increased risk of venous thromboembolism.^16^

The NOD-like receptor 3 (NLRP3) inflammasome has emerged as a central mediator of inflammation-associated vascular risk.^17,18^ Among its downstream effectors, interleukin (IL)-6 is a strong predictor of morbidity and mortality in ART-treated PWH and is consistently associated with cardiovascular and other non-AIDS events.^10,12,13,19–23^ Genetic Mendelian randomization (MR)^24^ studies support a causal role for IL-6 signaling in vascular disease in the general population,^25^ with emerging evidence in PWH.^26,27^

Upstream of IL-6, IL-1β is a key regulator of inflammasome-driven inflammation.^28^ Elevated IL-1β activity in monocytes is associated with increased IL-6 levels and systemic inflammation, implicating this axis in cardiovascular risk.^29^ Therapeutic targeting of IL-1β reduces vascular events in the general population^17,30^ without the off-target adverse lipid effects observed with IL-6 blockade,^17,30,31^ and and lowers inflammatory markers in PWH on ART,^32^ supporting a model in which NLRP3-mediated IL-1β and IL-6 signaling contributes to adverse cardiovascular outcomes.^33^

However, large-scale genomic studies of plasma IL-1β as a risk factor for chronic inflammatory diseases have been limited by difficulties in detecting its low circulating levels. Consequently, prior work has often relied on IL-18 as a surrogate, despite distinct biological functions.^34^ Recent high sensitivity assays now enable reliable quantification of IL-1β in ART-suppressed PWH,^35–37^ permitting epidemiologic exploration of upstream regulators of NRLP3 inflammasome-mediated effects on atherosclerotic disease progression.

Here, we performed the largest genome-wide analysis of host genetic determinants of circulating NLRP3 inflammasome-associated cytokines (IL-1β, IL-6, and IL-18) using whole genome sequencing data from 1,000 ART-suppressed PWH in the U.S. Military HIV Natural History Study. In contrast to prior HIV genomic studies focused on viral control or disease progression,^38–50^ we investigate inherited drivers of chronic immune activation. Using an integrative analytic framework informed by prior work,^51^ we identified multiple novel loci and pathways linking inflammasome signaling to immune, antiviral, and vascular biology. These findings define genetic mechanisms underlying persistent inflammation and highlight potential therapeutic targets to mitigate excess cardiovascular risk in PWH and the general population.

## RESULTS

### Study participant characteristics

We analyzed 1000 PWH from the U.S. Military HIV Natural History Study (NHS), a prospective cohort of >6,600 participants followed from HIV acquisition through long-term ART^52^ (**Fig. 1A**). All participants had confirmed HIV-1 infection, sustained virologic suppression for ≥1 year on ART, and available paired plasma and peripheral blood mononuclear cell specimens. After quality control, genetic data from 993 individuals were included in downstream analyses. The cohort comprised primarily individuals of European (EUR) and African (AFR) ancestry (**Fig. 1B**; **Supplementary Fig. 1**; **Supplementary Table 1**). Principal component analysis (PCA) demonstrated concordance between genetic and reported ancestry (**Fig. 1C**).

**Fig. 1.**
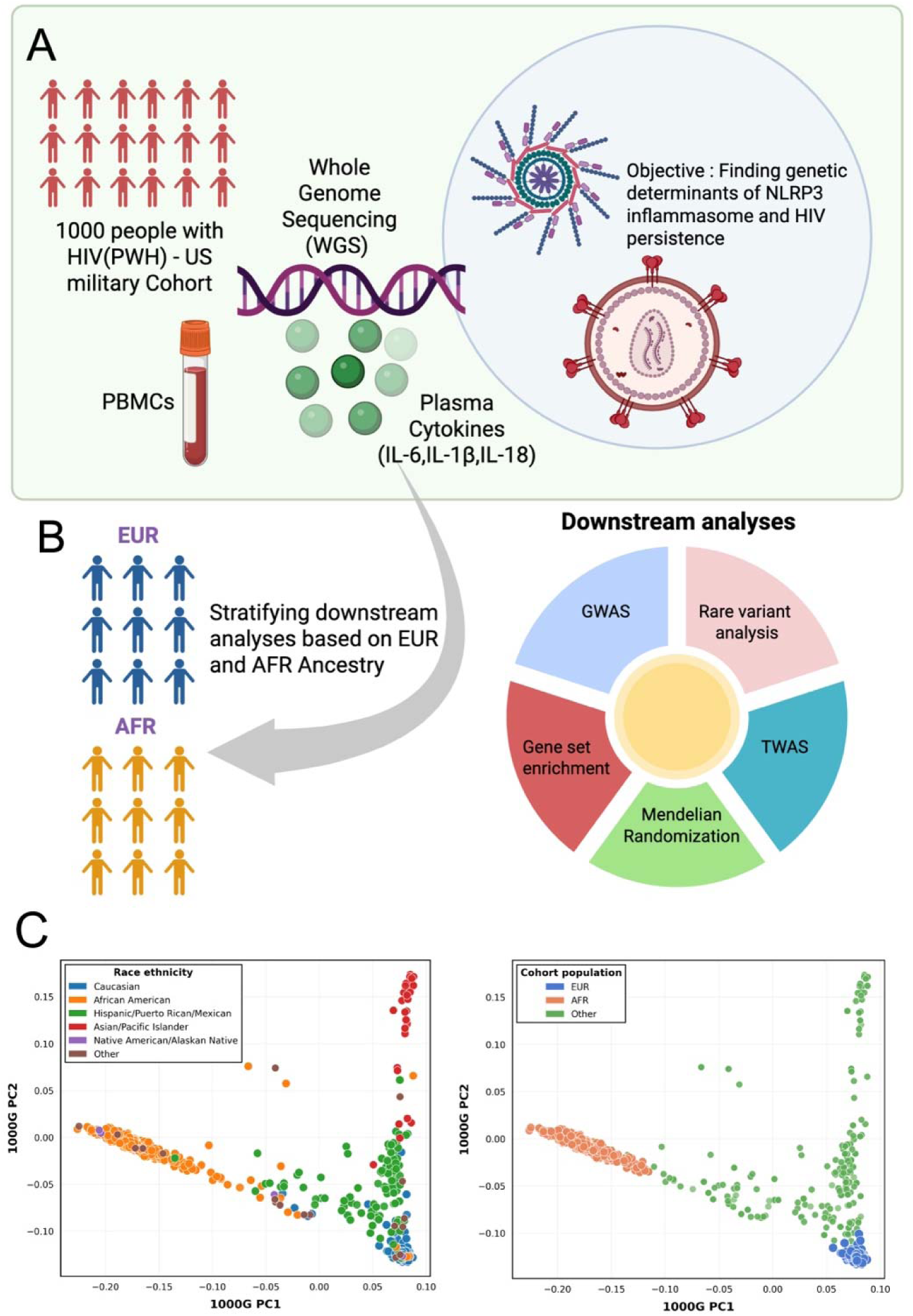
Study overview and population characteristics. Whole genome sequencing (WGS) was performed on 1,000 people with HIV (PWH) from the US Military HIV Natural History Study cohort, with plasma cytokines (IL-6, IL-1β, IL-18) and PBMCs collected to identify genetic determinants of NLRP3 inflammasome activation and HIV persistence (**A**). Participants were stratified by genetically inferred ancestry into European (EUR) and African (AFR) ancestry groups for downstream analyses including genome-wide association studies (GWAS), transcriptome-wide association studies (TWAS), rare variant analysis, gene set enrichment, and Mendelian randomization (**B**). Principal component analysis (PCA) of participant genotypes projected onto 1000 Genomes Project reference populations, colored by race ethnicity (left) and assigned ancestry based of 1000 genomes PCs (right), demonstrates broad concordance between genetic and self-reported ancestry (**C**).

### Differential quantifiability and variability of IL-6, IL-1**β**, and IL-18 in plasma from study participants

Plasma IL-6, IL-1β, and IL-18 were quantified in 1,000 ART-suppressed participants using a custom MSD U-Plex platform. Detectability was high (IL-18: 99.02%; IL-6: 97.28%; IL-1β: 91.81%), but quantifiability above the assay-specific limit of detection differed: IL-18 (97.24%; LOD = 0.50 pg/mL), IL-6 (82.14%; LOD = 0.33 pg/mL), and IL-1β (74.97%; LOD = 0.15 pg/mL) (**Supplementary Table 2**). Variability among quantifiable samples was lower for IL-18 (CV = 63.10%) than for IL-6 (1224.93%) or IL-1β (973.58%), consistent with pronounced right-skew for IL-6 and IL-1β.

Consistent with prior reports,^53,54^ IL-1β showed lower quantifiability than IL-6 and IL-18. Cytokine levels were log-transformed for downstream analyses. Although samples were processed in two batches, signal distributions varied across plates (**Supplementary Fig. 2A**). Plates with >2 of 8 standards exceeding two median absolute deviations were classified as anomalous, and ComBat correction^55^ was applied after log transformation. Post-correction distributions were well aligned (**Supplementary Fig. 2B-C**), indicating effective batch correction.

### Single-variant GWAS of NLRP3-inflammasome cytokines identified loci linked to atherogenesis and antiviral responses

After quality control, 66,084,459 variants from 993 participants were included in analyses (**Supplementary Fig. 1**). Additional filtering for ancestry outliers and relatedness yielded 417 EUR and 387 AFR participants. Genomic inflation (λ_GC_)^56^ was minimal (λGC=0.99±0.0087), indicating adequate control for population stratification (**Supplementary Fig. 3**).

Across six ancestry-stratified GWAS of IL-6, IL-1β, and IL-18, we identified 14 loci reaching genome-wide significance (p<5e-08) (**Supplementary Table 3**). One lead signal and two additional variants (p<1e-04) colocalized with GTEx^57^ expression quantitative trait loci (eQTLs), all showing effects in coronary artery tissue (**Supplementary Table 4**), supporting a role in vascular inflammatory pathways.

For IL-6, ancestry-specific associations were observed. In EUR, loci near *KLHL29* (previously associated with heart disease^58^) and *GOLGA6L1*, a Golgi-associated gene potentially involved in intracellular trafficking and immune regulatory processes^59,60^ (**Fig. 2A**; **Supplementary Fig. 4**). In AFR, six loci were detected, including *EPCAM, AGMO, EEA1, FGF9*, and *FTO/IRX3* (**Fig. 2A**; **Supplementary Fig. 5**). Notably, *EEA1*, involved in endocytosis and viral trafficking, has been implicated in HIV-1 replication and cell-to-cell transfer,^61,62^ suggesting a potential link between host genetics, viral processing, and immune activation. AGMO, a lipid-metabolizing enzyme regulating platelet-activating factor, has been associated with inflammatory signaling, including IL-1β expression in macrophages.^63,64^ The *AGMO* locus is distinct from a nearby IL-1β-associated signal at DGKB, indicating independent effects. Both loci have also been implicated in type 2 diabetes across ancestries.^65,66^

**Fig. 2.**
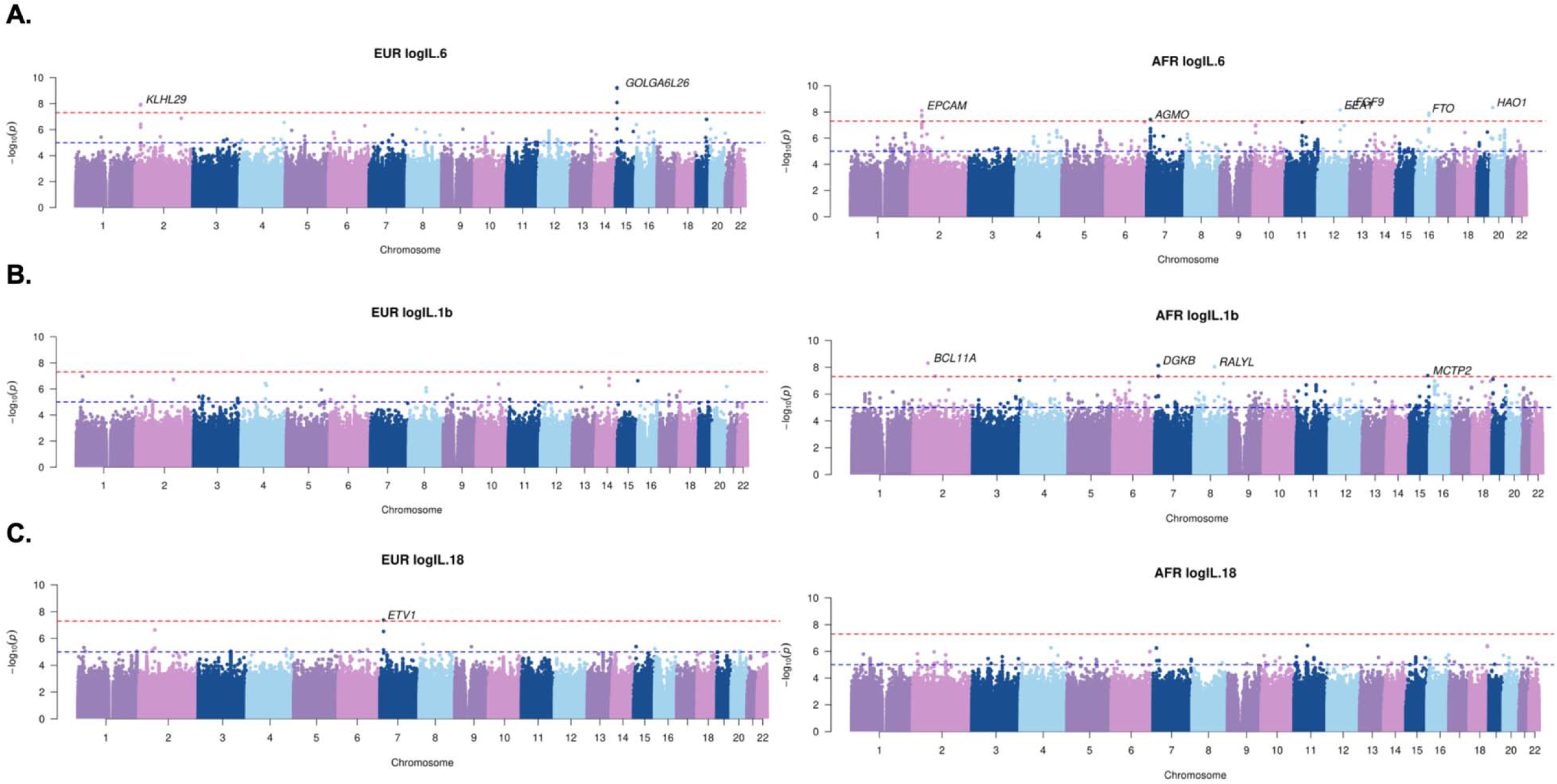
Manhattan plots of single-variant GWAS for NLRP3-inflammasome-associated cytokines. Genome-wide association analyses were performed for plasma levels of IL-6 (**A**), IL-1β (**B**), and IL-18 (**C**). Panels show results separately for participants of European ancestry (EUR; left) and African ancestry (AFR; right). Each point represents a single genetic variant, plotted by chromosomal position on the x-axis and -log_₁₀_(p-value) on the y-axis. The red horizontal line indicates the genome-wide significance threshold (p < 5e-08). Notable loci exceeding this threshold are highlighted in each plot. Differences in association patterns between EUR and AFR populations reflect both allele frequency differences and population-specific genetic architecture, illustrating the importance of ancestry-stratified analyses.

For IL-1β, four loci were identified in AFR (**Fig. 2B**; **Supplementary Fig. 6**), including *BCL11A*, a transcription factor implicated in HIV-1 replication.^67^ For IL-18, a genome-wide significant association with *ETV1* was observed in EUR (**Fig. 2C**; **Supplementary Fig. 7**). ETV1, together with fibroblast growth factor signaling (e.g., FGF9), has been linked to fibroblast activation and vascular remodeling.^68,69^ A related signal involving *FGF9* was observed for IL-6 in AFR (**Supplementary Table 3**).

Gene set enrichment analyses (GSEA) highlighted shared and ancestry-specific pathways. IL-6 loci were enriched for immune activation, autophagy, lipid metabolism, and vascular remodeling in EUR, and cell-cycle and intracellular signaling pathways in AFR (**Supplementary Table 5**); IL-1β loci were enriched for tissue remodeling and angiogenesis in EUR and immune and lipid pathways in AFR (**Supplementary Table 6**). IL-18 loci were enriched for DNA damage response and immune signaling in EUR and morphogenesis and growth factor signaling pathways in AFR (**Supplementary Table 7**).

### Gene-based rare variant analyses revealed immune, antiviral, and cardiovascular pathways

Gene-based rare variant analyses using *SMMAT* in GMMAT^70^ aggregated predicted functional variants (including high-impact and REVEL >0.6 missense variants) to test associations with IL-6, IL-1β, and IL-18. We identified 26 genes meeting the gene-based significance threshold (p<1/number of genes tested) (**Table 1**). Quantile-quantile plots showed modest inflation (**Supplementary Fig. 8**), likely reflecting the limited number of genes analyzed.

**Table 1.** Gene-based rare variant associations with NLRP3-inflammasome cytokines. Analyses were performed for plasma IL-6, IL-1β, and IL-18 in European (n=654) and African (n=616) ancestry participants using the Set Mixed Model Association Test (SMMAT) in the Generalized linear Mixed Model Association Tests (GMMAT) framework. Models were adjusted for age, sex, five genetic principal components, and genetic relatedness. The table includes genes meeting cytokine-specific significance, defined as the inverse of the number of genes tested for each cytokine within each ancestry. Top variants were annotated with cis-expression quantitative trait loci (cis-eQTLs) from GTEx for functional context.

| Gene | Gene Name | P-value | Relevance to vascular disease and/or HIV |
| --- | --- | --- | --- |
| **IL-6 (EUR)** |  |  |  |
| *ATP2C2* | ATPase Secretory Pathway Ca2+ Transporting 2 | 1.10e-03 | Involved in calcium signaling and potentially in arrhythmias such as atrioventricular nodal reentrant tachycardia<sup>114</sup> |
| *ACACB* | Acetyl-CoA Carboxylase Beta | 1.11e-03 | Associated with vascular endothelial cell senescence<sup>109</sup> |
| *COL21A1* | Collagen Type XXI Alpha 1 Chain | 1.13e-03 | Variant in *COL21A1* found to be associated with hypertension<sup>115</sup> |
| *FAT2* | FAT Atypical Cadherin 2 | 3.23e-05 | None noted |
| *SFXN1* | Sideroflexin 1 | 4.70e-04 | Involved in mitochondrial function, relevant to both metabolic and cardiovascular health<sup>116</sup> |
| **IL-6 (AFR)** |  |  |  |
| *CCDC168* | Coiled-Coil Domain-Containing Protein 168 | 4.20e-04 | None noted |
| **IL-1β (EUR)** |  |  |  |
| *RNF213* | Ring Finger Protein 213 | 1.03e-03 | Involved in blood vessel formation<sup>74</sup> |
| *NPHP4* | Nephrocystin 4 | 1.14e-03 | None noted |
| *CAPN1* | Calpain 1 | 1.26e-04 | Associated with atherosclerotic plaque and calcification<sup>108</sup> |
| *MYH13* | Myosin Heavy Chain 13 | 1.60e-04 | Involved in skeletal muscle contraction<sup>117</sup> |
| *LMNA* | Lamin A/C | 1.68e-04 | Associated with various cardiovascular diseases, including cardiomyopathy<sup>76</sup> |
| *TGFBR2* | TGF-β Receptor Type II | 2.82e-04 | TGF-β expression is associated with myocardial fibrosis in people with HIV<sup>110</sup> |
| *ABCA1* | ATP Binding Cassette Subfamily A Member 1 | 3.44e-04 | Involved in cholesterol transport and immune cell function<sup>72</sup> |
| *TFRC* | Transferrin Receptor | 5.19e-04 | Important for iron uptake and involved in pathogenesis of cardiovascular diseases<sup>91</sup> |
| *HNRNPH3* | Heterogenous Nuclear Ribonucleoprotein H3 | 5.61e-04 | Involved in RNA processing and regulation, potentially impacting immune response<sup>118</sup> |
| *ACACB* | Acetyl-CoA Carboxylase Beta | 6.73e-04 | See above |
| *EMP3* | Epithelial Membrane Protein 3 | 6.88e-04 | Involved in cell signaling and potentially in immune response<sup>119</sup> |
| *DST* | Dystonin | 9.23e-06 | None noted |
| **IL-1β (AFR)** |  |  |  |
| *NNT* | Nicotinamide Nucleotide Transhydrogenase | 1.35e-04 | Loss associated with increased production of vascular reactive oxygen species and development of atherosclerotic plaque<sup>95</sup> |
| *AGA* | Aspartylglucosaminidase | 1.38e-05 | Primarily known for its role in aspartylglycosaminuria, a lysosomal storage disease<sup>120</sup> |
| *GRAMD1A* | GRAM Domain Containing 1A | 1.53e-04 | Involved in lipid transfer and potentially cell signaling<sup>121</sup> |
| *ABCA12* | ATP Binding Cassette Subfamily A Member 12 | 2.17e-06 | Important in cholesterol efflux in macrophages, a buildup of cholesterol can lead to atherosclerosis<sup>72</sup> |
| *OGFOD2* | 2-Oxoglutarate and Iron Dependent Oxygenase Domain Containing 2 | 2.96e-05 | None noted |
| *ABCC1* | ATP Binding Cassette Subfamily C Member 1 | 8.60e-05 | Involved in transport of HIV protease inhibitors<sup>73</sup> |
| **IL-18 (EUR)** |  |  |  |
| *DDX17* | DEAD-Box Helicase 17 | 3.87e-04 | Necessary for HIV production<sup>77</sup> |
| *DDX41* | DEAD-Box Helicase 41 | 7.57e-04 | Involved in retrovirus reverse transcription<sup>78</sup> |
| *TAOK2* | TAO Kinase 2 | 9.67e-05 | Involved in MAPK signaling, which has roles in both immune response and cellular stress<sup>94</sup> |

For IL-1β, significant associations included multiple ATP-binding cassette (ABC) transporter genes (*ABCA12, ABCC1*, and *ABCA1*), which regulate lipid transport and macrophage cholesterol efflux.^71^ Dysregulation of these pathways promotes foam cell formation and atherogenesis,^72^ suggesting a genetic link between IL-1β-mediated inflammation and lipid-driven vascular disease. *ABCC1*, which transports multiple xenobiotics including antiretroviral agents,^73^ may additionally influence intracellular drug exposure and inflammatory signaling. Rare variant associations were also identified in genes related to vascular remodeling and cardiac structure. *RNF213*, which has been implicated in angiogenesis and vascular disease,^74^ and is responsive to inflammatory signaling,^75^ was associated with cytokine levels. *LMNA*, which encodes nuclear lamins involved in nuclear integrity and cardiomyopathy,^76^ was associated with IL-1β in AFR participants, linking structural and inflammatory pathways.

Gene-based analyses further highlighted antiviral and innate immune pathways, particularly for IL-18. Two DEAD-box helicases, *DDX17* and *DDX41*, were associated in EUR participants; these genes regulate RNA metabolism and innate immune sensing and have established roles in HIV replication and host antiviral responses.^77,78^

Gene set enrichment analyses supported these findings, revealing pathways related to immune activation, viral replication, cardiovascular signaling, and metabolism (**Fig. 3**). Enrichment patterns were ancestry-specific. For IL-6, EUR-associated genes were enriched for antiviral and immune pathways, including type I interferon (IFN-I) signaling, chemokine signaling (including CXCR4/CXCR5), and epidermal growth factor (EGF) signaling, whereas AFR-associated genes were enriched for metabolic and vascular pathways, including mTOR^79^ signaling and endothelial cell migration (**Supplementary Table 8**). For IL-1β, EUR-associated genes were enriched for inflammatory pathways, while AFR-associated genes showed broader enrichment across innate and adaptive immune signaling, including Toll-like receptor, IFN-I/II, T- and B-cell receptor pathways, as well as angiogenesis (**Supplementary Table 9**). For IL-18, EUR-associated genes were enriched for natural killer (NK) cell activity, antigen presentation, TGF-β and p38 MAPK signaling, and epigenetic regulation, whereas AFR-associated genes were enriched for pathways related to tissue remodeling, MAPK activation, IL-4 signaling, and coagulation (**Supplementary Table 10**).

**Fig. 3.**
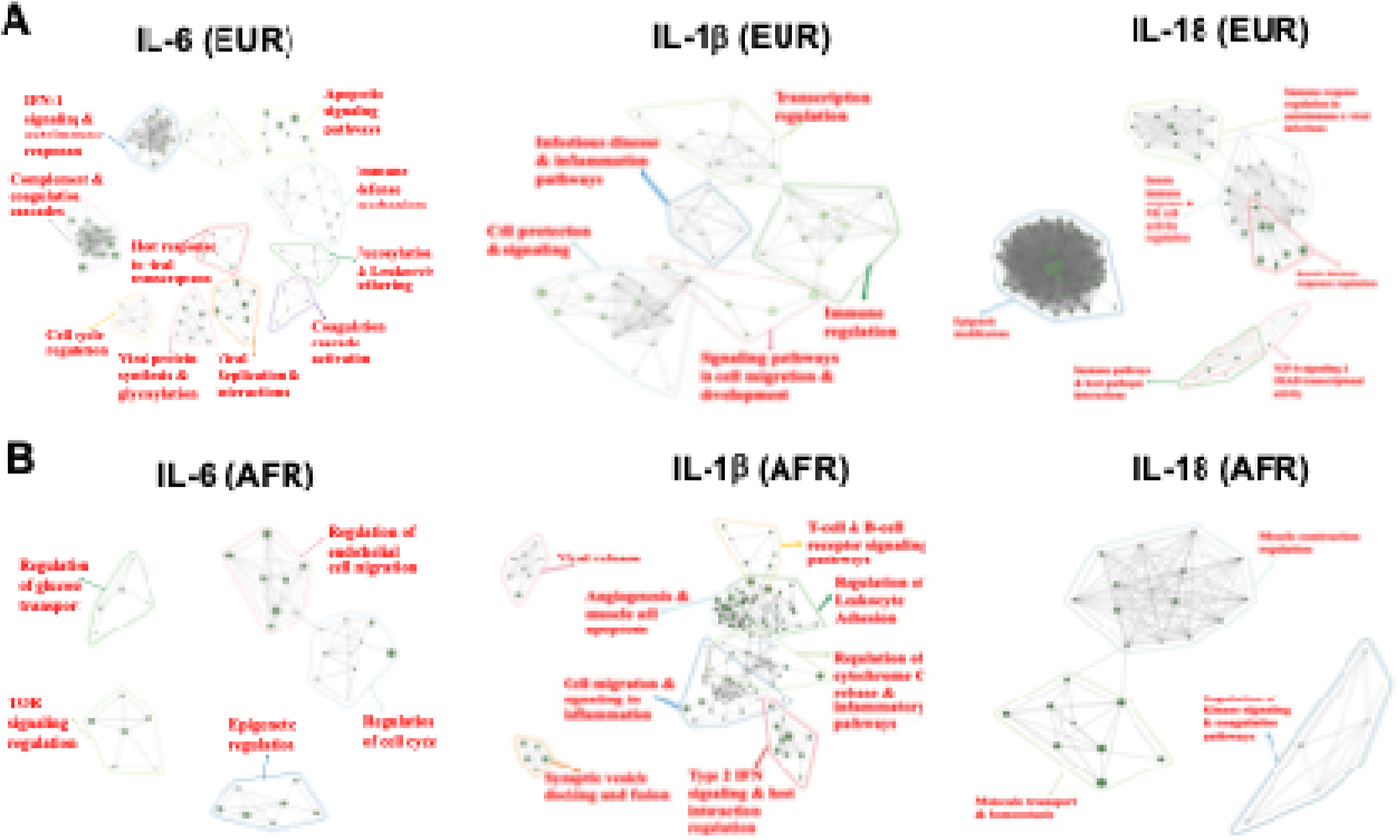
Gene set enrichment analysis (GSEA) of gene-based rare variant associations for NLRP3-inflammasome–associated cytokines. Gene-based rare variant associations were analyzed separately in participants of European (EUR; **A**) and African (AFR; **B**) ancestry for plasma IL-6, IL-1β, and IL-18. Enrichment analysis identifies biological pathways overrepresented among associated genes. Data are visualized as a network of nodes representing gene sets, linked based on shared genes, with clusters manually annotated. GSEA was performed using MSigDB Hallmark, Gene Ontology, and Reactome gene sets. Pathways with p < 0.05 were selected and clustered using the Jaccard index (threshold 0.2) via vissE software. Node size indicates the number of genes in each set, and edges represent the degree of overlap between gene sets based on Jaccard index values. Each label represents a biologically coherent group of pathways and was assigned based on the top representative pathways within each cluster.

### Transcriptome-wide association study identified immune and tissue-specific regulators of inflammasome cytokines

To assess whether genetically regulated gene expression influences NLRP3 inflammasome-associated cytokine levels, we performed transcriptome-wide association studies (TWAS) across four relevant tissues (coronary artery, left ventricle, spleen, and whole blood) in AFR and EUR cohorts. We identified 18 significant TWAS signals across 17 genes (p<2e-04) without evidence of inflation (**Fig. 4**; **Supplementary Table 11**; **Supplementary Fig. 9**). The strongest association was observed for *INPP5J* in coronary artery tissue in AFR participants, where higher predicted expression was associated with increased IL-1β (p=4.8e-08). INPP5J belongs to a phosphatase family that includes regulators of NLRP3 inflammasome signaling.^80^ In cardiac tissue, *PDCD6* expression was positively associated with IL-6 in AFR participants, whereas *TGFB1* expression was inversely associated with IL-18 in EUR. PDCD6 participates in ESCRT-mediated viral budding,^81^ suggesting a link between host gene regulation, HIV replication, and immune activation.

**Fig. 4.**
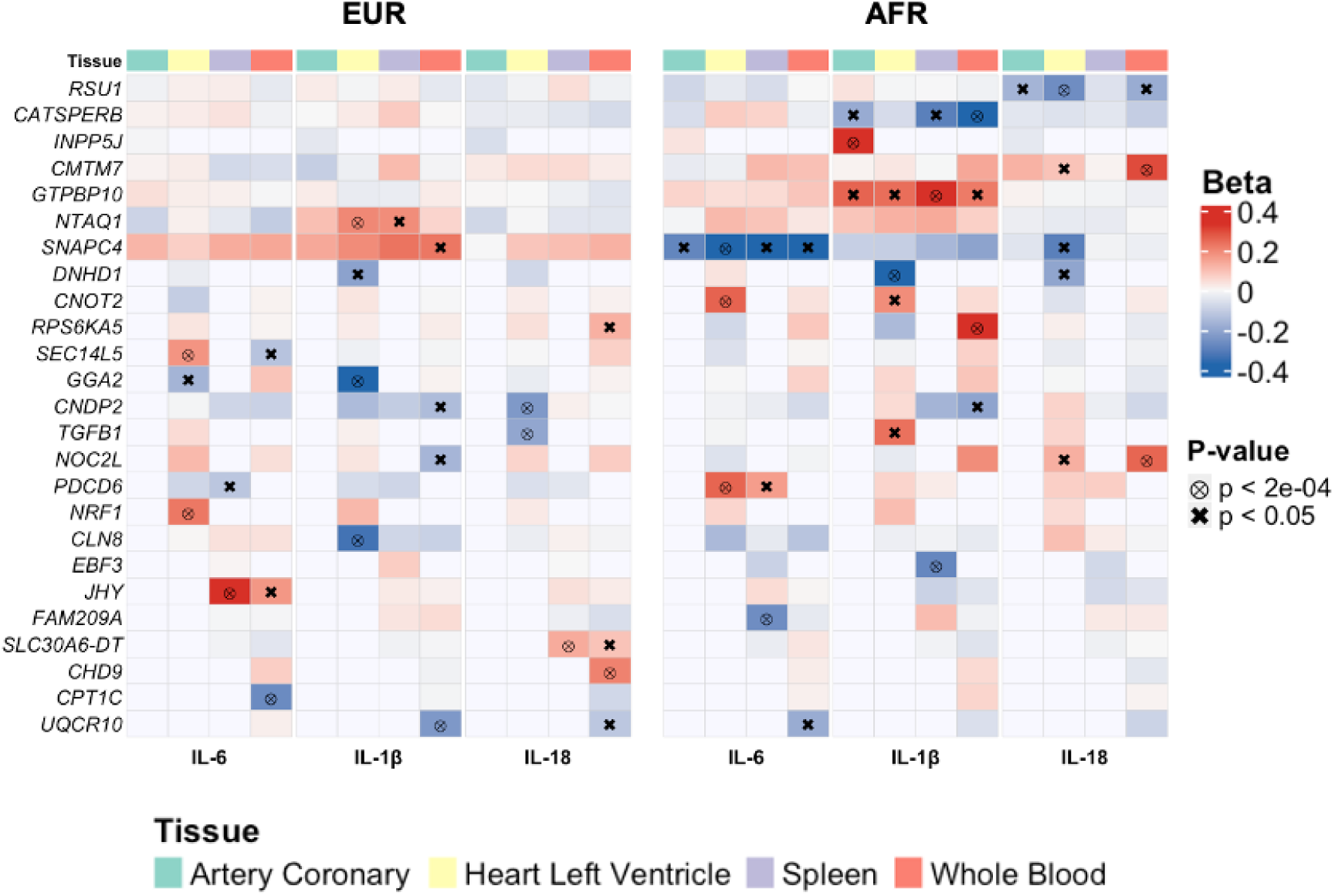
Heatmap of top gene hits from transcriptome-wide association study (TWAS) of NLRP3-inflammasome cytokines. Gene-based associations with plasma IL-6, IL-1β, and IL-18 were identified using predicted gene expression in four GTEx tissues. Associations were selected using a p-value threshold of 1/n, where n is the number of genes tested for each tissue–cytokine–ancestry combination. The heatmap shows genes on the y-axis, with cytokines on the x-axis. Tissues are indicated in the top row by color: green = coronary artery, yellow = heart left ventricle, lavender = spleen, and peach = whole blood. The left side of the heatmap shows European ancestry (EUR) participants, and the right side shows African ancestry (AFR) participants. Beta values represent effect sizes, and symbols indicate significance: ⊗ denotes associations meeting the most stringent p-value threshold (p < 2e-04); × indicates p < 0.05.

Gene set enrichment analyses revealed tissue- and ancestry-specific pathways linking cytokine-associated gene expression to immune, antiviral, metabolic, and cardiovascular processes (**Fig. 5**; **Supplementary Tables 12-14**). In cardiovascular tissues, enriched pathways included vascular remodeling, endothelial migration, and signaling pathways such as IFN, AKT, VEGF, and integrin signaling. In immune-relevant tissues (spleen and whole blood), enriched pathways included type I interferon signaling, T-cell and myeloid activation, antiviral responses (including ISG15 signaling), and lipid metabolism.

**Fig. 5.**
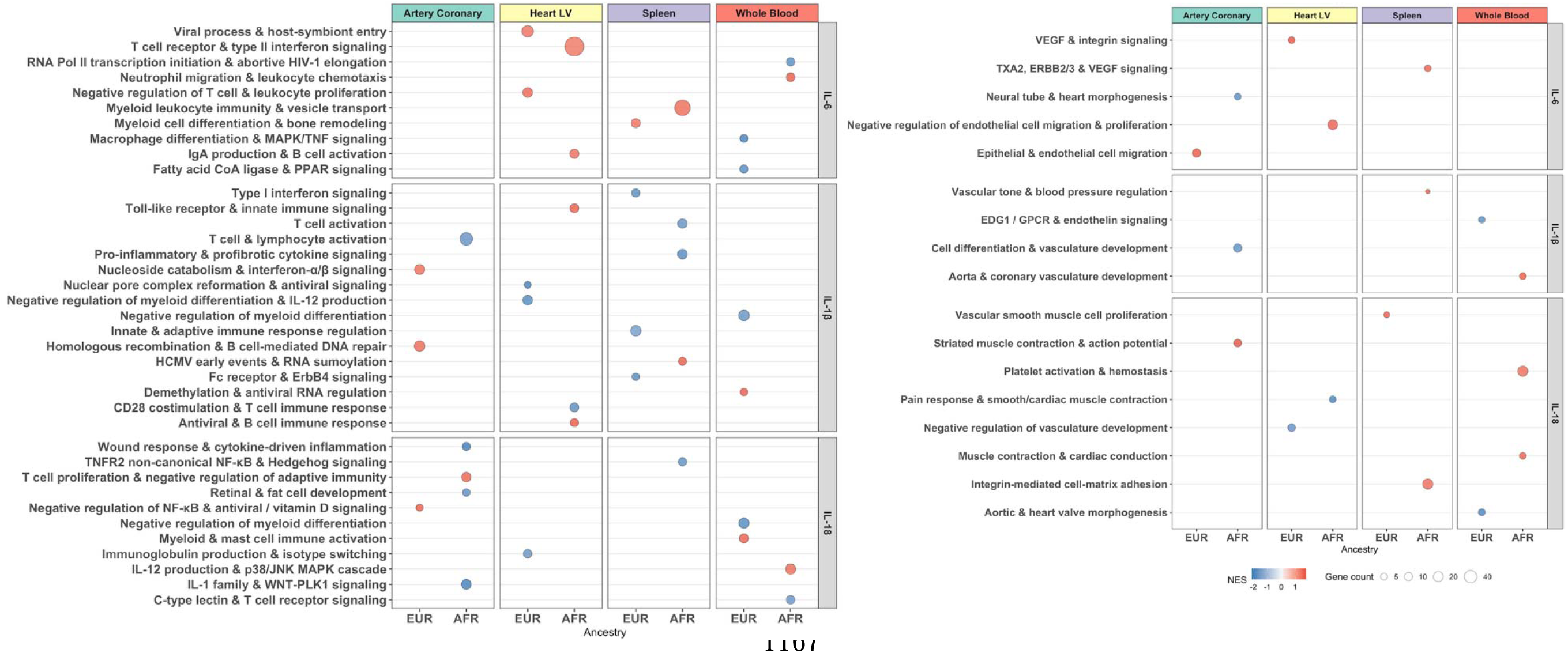
Gene set enrichment analysis (GSEA) identifying pathways associated with circulating NLRP3 inflammasome cytokine levels, grouped into Antiviral, Immune & Inflammatory Pathways (left) and Vascular, Cardiac & Homeostasis Pathways (right). GSEA was performed on TWAS results using Hallmark, Gene Ontology, and Reactome gene sets, and results are shown faceted by cytokine (rows: IL-6, IL-1β, IL-18) and tissue (columns: Artery Coronary, Heart Left Ventricle [Heart LV], Spleen, Whole Blood), with ancestry (EUR, AFR) on the x-axis. Enriched pathways were clustered based on overlap using a Jaccard index threshold of 0.2. The y-axis displays cluster labels, where each label represents a biologically coherent group of pathways and was assigned based on the top representative pathways within each cluster; all constituent pathways for each cluster are provided in Supplementary Tables 11–13. Circle size reflects the number of core genes contributing to each gene set. Gene sets with increased predicted transcription are shown in red; those with decreased predicted transcription are shown in blue.

Across cytokines, IL-6 and IL-1β were associated with immune and vascular pathways, whereas IL-18 was enriched for inflammasome signaling, NK cell activation, and epigenetic regulation. Enrichment patterns differed by ancestry, with stronger inflammatory and vascular signals in AFR and greater antiviral and immune regulatory pathways in EUR.

### Mendelian randomization analyses causally linked inflammasome cytokines to vascular risk

We evaluated the causal effects of NLRP3-inflammasome–associated cytokines on vascular outcomes using Mendelian randomization.^24^ Atherosclerotic cardiovascular disease (ASCVD) was defined as incident coronary artery disease (CAD), myocardial infarction (MI), cerebrovascular accident (CVA), or peripheral arterial disease (PAD), while vascular events (VE) additionally included venous thrombotic events (VTE: deep vein thrombosis [DVT] and pulmonary embolism [PE]). After excluding prevalent and early events, analyses included 133 VE cases (including 110 ASCVD) and 694 controls (**Supplementary Fig. 10**). Cases and controls were broadly balanced for demographic and HIV-related characteristics, with a higher prevalence of cardiometabolic comorbidities among cases (**Supplementary Table 15**).

Disaggregated Mann-Whitney tests showed that IL-6 and IL-18 levels were elevated across ASCVD subtypes, with significant increases in CAD, whereas no consistent differences were observed for VTE (**Supplementary Fig. 11**). Across VE diagnoses, plasma IL-18 showed the most consistent elevation relative to controls.

MR analyses were performed separately by ancestry using cytokine-associated variants as instrumental variables. Models were adjusted for age, sex, and ancestry (principal components and genetic relatedness matrix), with consistent results across MR methods (IVW, MR-Egger, weighted median, MR-PRESSO) and minimal evidence of heterogeneity or directional pleiotropy (**Fig. 6**; **Supplementary Tables 16-18**).

**Fig. 6.**
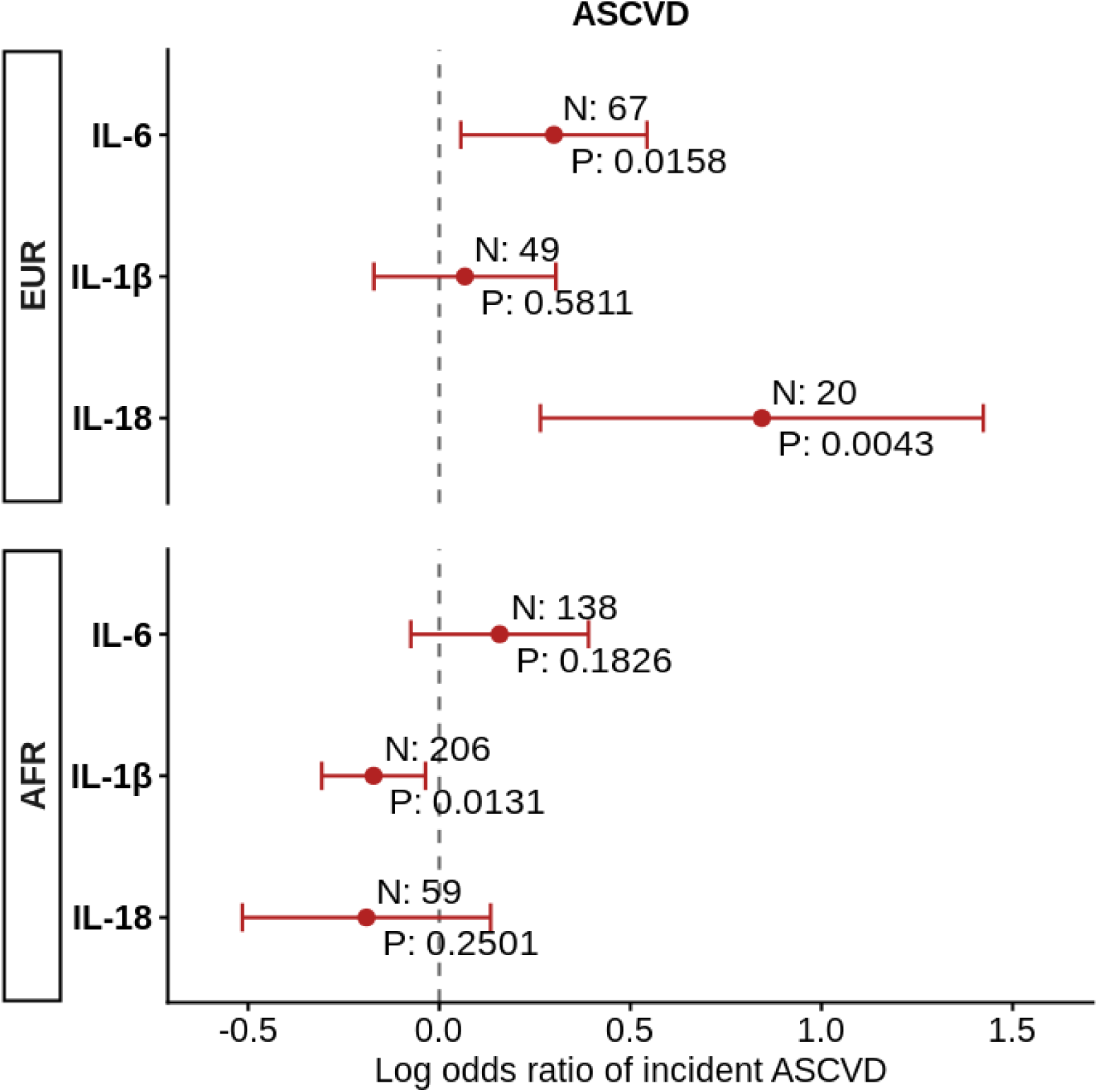
Forest plot of Mendelian randomization (MR) estimates of the causal effects of NLRP3 inflammasome–associated cytokines on incident atherosclerotic cardiovascular disease (ASCVD). Forest plots show inverse-variance weighted MR odds ratios (ORs) for incident ASCVD per genetically predicted increase in plasma IL-6, IL-1β, and IL-18 levels, using cytokine-associated genetic variants as instrumental variables. Results are presented separately for participants of European (EUR; top) and African (AFR; bottom) ancestry. Models were adjusted for age, sex, first five principal components (PCs). The x-axis denotes the log OR for ASCVD per two-fold increase in genetically predicted, log-transformed cytokine concentrations. N indicates the number of genetic variants comprising each cytokine-specific instrumental variable, selected based on association with plasma cytokine levels (p < 1e-05), minor allele frequency > 0.01, and instrument strength (F-statistic > 10). P denotes the p-value for the MR effect estimate.

In EUR participants, genetically predicted IL-6 and IL-18 levels were associated with increased ASCVD risk (IL-6: log OR = 0.30, 95% CI 0.06-0.54, p=1.6e-02; IL-18: log OR = 0.84, 95% CI 0.26-1.42, p=4.3e-03), while IL-1β showed a positive but non-significant association. In AFR participants, IL-6 showed a positive but non-significant association, whereas IL-1β was inversely associated with ASCVD risk (log OR = −0.17, 95% CI -0.31 to -0.04, p=1.3e-02), with a similar non-significant trend for IL-18. Analysis of MAF-stratified SNP effect estimates, as previously described,^82^ revealed no systematic patterns indicative of residual population stratification (**Supplementary Fig. 12**).

Single-variant analyses highlighted biologically relevant loci contributing to these associations, including variants near *NCK2*, which promotes endothelial inflammation and atherosclerosis progression,^83^ *CREB1*, a transcription factor involved in immune activation and HIV susceptibility,^84^ and *GPD2*, which regulates oxidative stress and platelet activation pathways linked to coagulation,^85^ all showing positive effects on ASCVD risk in EUR participants. In AFR participants, variants near *TMCC3*, implicated in fatty liver and metabolic dysfunction,^86^ and *MAT2B*, which regulates S-adenosylmethionine metabolism and HIV latency pathways,^87^ showed effect estimates consistent with inverse associations (**Supplementary Fig. 13**; **Table 2**).

**Table 2.** Selected SNPs with relevance to vascular disease and/or HIV from Mendelian randomization (MR) analysis of NLRP3-inflammasome cytokines and atherosclerotic cardiovascular disease (ASCVD). Shown are selected single-nucleotide polymorphisms (SNPs included in the multivariable MR analysis presented in **Fig. 6**), which estimates the effects of genetically predicted plasma cytokine levels (for example, IL-6) on risk of incident ASCVD while accounting for other cytokines within the NLRP3 inflammasome pathway (including IL-1β and IL-18). The SNPs listed are of particular relevance to vascular disease and/or HIV due to their established associations with cytokine regulation and clinical outcomes. Results are stratified by cytokine (IL-6, IL-1β, IL-18) and by ancestry (European [EUR] and African [AFR]). Columns report the SNP (chromosomal position and allele change), rsID, mapped gene, and effect estimates for each stage of the MR framework. Beta (cytokine) denotes the association between each SNP and plasma cytokine levels (natural log scale), with positive values indicating higher and negative values indicating lower cytokine concentrations. Beta (ASCVD) represents the association between each SNP and incident ASCVD risk, where positive values indicate increased risk and negative values indicate reduced risk. Beta (MR) reflects the estimated effect of plasma cytokine levels on ASCVD risk derived from the MR analysis, with positive values indicating higher cytokine levels associated with increased ASCVD risk and negative values indicating a protective association. Standard errors (SE) and p-values (P) are provided for the overall MR estimates.

| SNP | rsID | Gene Name | Beta (Cytokine) | Beta (ASCVD) | Beta (MR) | P | SE | Relevance to vascular disease and/or HIV |
| --- | --- | --- | --- | --- | --- | --- | --- | --- |
| IL-6 (EUR) |  |  |  |  |  |  |  |  |
| chr2_105839158_C_T | rs78576957 | NCK2 | -0.45 | 0.93 | 2.63 | 4.3e-02 | 1.30 | Promotes atherosclerosis progression in endothelium<sup>83</sup> |
| IL-6 (AFR) |  |  |  |  |  |  |  |  |
| IL-1β (EUR) | | | | | | | | |
| chr2_207601054_A_G | rs2551928 | CREB1 | 0.37 | 0.77 | 2.1 | 3.2e-02 | 0.98 | Associated with reduced risk of HIV acquisition and increased efficacy of vaccination<sup>84</sup> |
| IL-1β (AFR) | | | | | | | | |
| chr12_94741347_A_G | rs139897304 | TMCC3 | -1.34 | 1.80 | -1.34 | 1.3e-02 | 0.54 | High expression of *TMCC3* is associated with Non-alcoholic/alcoholic fatty liver disease, a risk factor for cardiovascular diseases<sup>86</sup> |
| chr5_163542203_A_G | rs10045036 | MAT2B | -0.74 | 1.42 | -1.93 | 2.8e-03 | 0.65 | Regulates S-adenosylmethionine (SAM) production by controlling *MAT2A*. HIV is suppressed via SAM-mediated epigenetic modifications<sup>87</sup> |
| chr12_81426959_T_C | rs540015898 | PPFIA2 | -1.59 | 1.70 | -1.07 | 4.9e-02 | 0.54 | Encodes for liprin-α1, which is implicated in endothelial cell migration<sup>122</sup> |
| IL-18 (EUR) |  |  |  |  |  |  |  |  |
| chr2_156463682_C_T | rs4664207 | GPD2 | 0.22 | 0.67 | 3.10 | 7.2e-03 | 1.15 | Associated with impaired coagulation, altered ROS/NF-kb signaling and changes in platelet activation<sup>85</sup> |
| IL-18 (AFR) |  |  |  |  |  |  |  |  |

### Integration with CRISPR-based functional datasets revealed links between cytokine-associated loci, HIV host dependency, and T cell gene regulation

To functionally contextualize cytokine-associated loci, we integrated genes identified from GWAS (p<1e-05), rare variants, and TWAS analyses with CRISPR-based functional datasets in primary human CD4⁺ T cells, including genome-wide CRISPRa/CRISPRn of HIV host dependency and host defense response pathways^88,89^ and Perturb-seq transcriptional profiling.^90^

At the gene set level, GWAS-identified genes for IL-1β and IL-6 were enriched among host defense factors identified in CRISPRa screen (IL-1β: OR = 2.35, p=1.9e-02; IL-6: OR = 2.21, p=4.7e-02), indicating that transcriptional activation of these genes reduced HIV infectivity more than expected by chance (**Supplementary Table 19**). A similar but weaker trend was observed when aggregating all cytokine-associated genes across analyses, whereas no enrichment was observed in CRISPRn screens.^89^

At the individual gene level, CRISPRa analyses identified both host defense and host proviral factors. *TFRC*, which regulates iron metabolism and has been linked to cardiovascular disease and NK cell activation^91^ and *CLN8*, which is known to play a role in lipid homeostasis and lysosomal enzyme trafficking,^92^ exhibited antiviral effects. In contrast, *CAPN1* (associated with HIV Tat-mediated neurotoxicity, neutrophil apoptosis, and atherosclerotic plaque formation),^93^ *FGF9* (linked to fibroblast activation and vascular remodeling),^68^ *TAOK2* (involved in MAPK signaling),^94^ *NNT* (associated with oxidative stress and atherosclerosis)^95^ and *SLC30A6* (linked to zinc-dependent antiviral responses)^96^ exhibited proviral activity, with increased HIV infectivity upon overexpression (FDR<0.2) (**Supplementary Fig. 14**). Directionally consistent but weaker effects were observed in CRISPRn screens.

Perturb-seq analyses further supported regulatory links between candidate genes and cytokine signaling. In the upstream direction, IL-6-associated genes were enriched among regulators of IL6 expression (OR = 3.36, p=7.1e-03), with knockdown of *JHY* and *RSU1*, associated with vascular inflammation and recurrence of ischemic stroke,^97^ increasing *IL6* expression. Additional cytokine-specific effects included *IRX3* (promotes angiogenesis by mediating endothelial cell migration^98^) knockdown, reducing *IL1R1* expression, and multiple genes involved in immune signaling, metabolic regulation, and vascular remodeling (*TGFBR2, CREB1, RPS6KA5, ACACB, FTO*), altering *IL18R1* expression across stimulation conditions (**Supplementary Tables 20-21**). In the downstream direction, cytokine receptor perturbations altered expression of multiple candidate genes, including *EMP3, TGFBR2, HNRNPH3, RSU1, LMNA*, *DDX17*, and *KLHL29*, indicating reciprocal regulatory interactions between cytokine signaling and candidate loci involved in immune regulation, RNA processing, endothelial/vascular biology, and HIV-related host functions. Together, these analyses provide functional support for cytokine-associated loci, linking genetic signals to HIV infectivity and transcriptional regulation in CD4⁺ T cells.

### Cross-cohort validation of IL-6 genetic associations in external population datasets

To assess generalizability of IL-6 GWAS findings, we evaluated external cohorts from the All of Us Research Program^99^ (N = 570), stratified by ancestry (**Supplementary Table 22**; **Supplementary Fig. 15A**). Genome-wide analyses showed minimal inflation (λ_GC_=1.0075 EUR; 1.0253 AFR) (**Supplementary Table 23**; **Supplementary Fig. 15B**) and identified nominal associations near genes involved in immune and vascular biology, including *ABCA1*, which regulates cholesterol efflux and macrophage lipid homeostasis^100^ (**Supplementary Fig. 15C-D**). Variants showing suggestive association in the discovery cohort showed no excess overlap with nominally associated variants in the validation cohorts in either EUR or AFR strata (**Supplementary Table 24**), with approximately equal proportions of concordant and discordant effect directions.

In contrast, pathway-level analyses demonstrated substantial cross-cohort consistency. Normalized enrichment scores were strongly correlated between the discovery and validation cohorts in both EUR (Spearman R=0.716, P=3.22e-47) and AFR (R=0.563, p=1.05e-25). Shared pathways converged on immune and vascular regulatory programs, including TGF-β (context-dependent role in ASCVD^101^), IL7, a key regulator of T-cell homeostasis and chronic immune activation,^102^ and Rap1 signaling, which is important for endothelial barrier function and vascular homeostasis^103^ (**Supplementary Tables 25-26**).

## DISCUSSION

In this study, we integrated whole genome sequencing, cytokine profiling, and longitudinal clinical data in ART-suppressed people with HIV to define the genetic architecture of NLRP3 inflammasome–associated cytokines and their relationship to vascular risk. By combining single-variant, rare variant, and transcriptome-wide association analyses with functional validation using recent genome-wide CRISPR-based perturbation datasets in primary human CD4⁺ T cells, we provide convergent evidence linking genetic variation to biological mechanisms. Across these complementary approaches, we identify coordinated genetic regulation connecting immune activation, antiviral defense, lipid metabolism, and vascular remodeling to circulating IL-6, IL-1β, and IL-18 levels and downstream atherosclerotic cardiovascular disease.

Prior host genetic studies in HIV have largely focused on virologic phenotypes, including viral control and reservoir size, with reproducible associations in the HLA region and *CCR5*.^38–45,47,104,105^ In contrast, comparatively few studies have examined genetic predictors of immunologic biomarkers, particularly in virally-suppressed PWH where residual inflammation persists despite ART.^51^ Our findings extend this framework by demonstrating that variation in inflammasome-associated cytokines is shaped by host genetic pathways that intersect antiviral defense, immune signaling, and cardiovascular biology, supporting the concept that persistent inflammation in treated HIV infection is, in part, genetically determined.

Across GWAS, rare variant, and TWAS analyses, multiple lines of evidence converged on pathways linking viral trafficking, lipid metabolism, macrophage recruitment, atherosclerosis, and NLRP3 inflammasome activation. Several loci implicated host-virus interactions, including *EEA1* (regulates endosomal trafficking and HIV entry^61,62^), *PDCD6* (involved in HIV virion release^81^), and DEAD-box helicases, *DDX17* and *DDX41* (required for HIV replication and innate immune sensing^77,78^). These findings suggested that genetic regulation of viral processing and sensing pathways may influence inflammasome-associated cytokine responses. In parallel, several loci pointed to a link between lipid handling (*AGMO*^63^), cholesterol efflux *(ABCA1, ABCA12, ABCC1*)^71–73^, vascular remodeling (F*TO/IRX3*^106^*, RNF213*^74^*, LMNA*^76^), macrophage recruitment and atherosclerosis (*ETV1*^68,69^), and NLRP3 inflammasome regulation (*INPP5J*^80^).

TWAS analyses provided additional insight into tissue-specific regulatory mechanisms. Associations in coronary artery and cardiac tissues highlighted pathways related to endothelial migration, vascular remodeling, and growth factor signaling, whereas immune tissues showed enrichment for interferon signaling, T cell activation, and antiviral responses. Notably, *INPP5J*, which has been implicated in the regulation of NLRP3 inflammasome signaling,^80^ and *PDCD6*, which participates in HIV virion release,^81^ linked tissue-specific gene expression with both inflammatory signaling and viral persistence. These findings supported a model in which genetically regulated expression programs across immune and vascular tissues contributed to inter-individual variation in circulating cytokine levels.

Mendelian randomization analyses suggested that genetically influenced variation in inflammasome-associated cytokines contributed to ASCVD risk in PWH. In EUR participants, IL-6 and IL-18 were associated with increased ASCVD risk, consistent with extensive epidemiologic and interventional evidence implicating IL-6 signaling in atherogenesis.^10,12,13,17–23,25–28^ In contrast, IL-1β-associated variants in AFR participants showed inverse associations with ASCVD, with similar trends for IL-18. Examination of individual SNPs contributing to these MR instruments identified biologically plausible loci whose known functions were directionally consistent with the observed associations. In EUR participants, IL-6-associated variants near genes implicated in endothelial inflammation and atherosclerosis progression were associated with increased ASCVD risk. In AFR participants, variants near genes involved in lipid metabolism, inflammatory signaling, and intracellular trafficking showed effect estimates consistent with the inverse MR associations, raising the possibility that genetic context may modify the relationship between inflammasome signaling and vascular risk. Although these findings required cautious interpretation given limited statistical power, they suggested potential heterogeneity in inflammasome-associated vascular risk across ancestry groups.

Functional integration with CRISPR-based perturbation datasets provided orthogonal support for these genetic findings and refined biological interpretation. Cytokine-associated loci were enriched among genes involved in host defense responses and pathways related to HIV persistence and chronic immune activation. At the gene level, loci related to oxidative stress and mitochondrial function (*NNT*^95^), iron metabolism (*TFRC*^91^), and MAPK signaling (*TAOK2*^94^) demonstrated effects on HIV infectivity, while genes such as *SLC30A6*, which has been linked to inhibition of HIV protease and reverse transcriptase activity,^96^ and *CAPN1*, which has been associated with HIV Tat-mediated neurotoxicity, neutrophil apoptosis, and atherosclerotic plaque formation,^93,107,108^ showed proviral activity. Perturb-seq analyses further identified bidirectional regulatory relationships between candidate genes and cytokine signaling pathways, including genes involved in endothelial function (*RSU1*^97^, *IRX3*^98^, *ACACB*^109^) and immune regulation (*TGFBR2* ^110^, *CREB1*^84^), suggesting that inflammasome-associated cytokines participated in feedback networks linking immune activation and vascular remodeling.

Comparisons with external population-based cohorts showed limited concordance at the individual variant level, but given differences in HIV status, ancestry composition, and sample size across cohorts, these analyses were interpreted as exploratory assessments of biological generalizability rather than strict variant-level replication.^111^ Nonetheless, pathway-level enrichment analyses demonstrated strong cross-cohort consistency in both EUR and AFR populations, including shared pathways related to immune regulation such as TGF-β signaling, which plays a context-dependent role in ASCVD,^101^ IL7, a key regulator of T-cell homeostasis and chronic immune activation,^102^ and Rap1 signaling, which regulates endothelial barrier function and vascular homeostasis.^103^ This suggests that although HIV-associated inflammation may alter detectable variants, broader IL-6 regulatory pathways remain partially conserved across populations.

This study had several strengths, including whole genome sequencing, high-sensitivity cytokine quantification, and longitudinal adjudication of vascular outcomes in a deeply phenotyped cohort. Integration of multiple genomic and functional approaches enabled identification of convergent biological pathways and provided complementary evidence for causal relationships.

Several limitations should be considered. Circulating cytokine levels may not fully capture cell-type-specific inflammasome activity, and the modest number of vascular events limited power for causal inference, particularly in ancestry-stratified analyses. Although multiple strategies were used to mitigate population stratification, including principal components, genetic relatedness modeling, and ancestry-stratified analyses, residual confounding could not be excluded.^112,113^ Functional validation relied on CD4⁺ T cell-based CRISPR systems, which may not fully reflect myeloid or vascular cell biology, and transcriptional readouts may not directly correspond to cytokine protein levels. In addition, external validation was limited to IL-6 in cohorts without HIV, which may restrict generalizability.

In summary, our findings demonstrated that both common and rare genetic variation shaped inflammasome-associated cytokine levels in PWH and linked these pathways to cardiovascular risk. These results highlighted the integration of antiviral defense, immune activation, and metabolic regulation in chronic inflammation, and suggested that ancestry-specific genetic effects may influence vascular risk. Future studies incorporating larger multi-ancestry cohorts, single-cell approaches, and functional genomics will be critical to refine these mechanisms and identify therapeutic targets to reduce inflammation-driven cardiovascular disease in PWH and the general population.

## METHODS

### Study participants

A total of 1000 PWH were selected from the U.S. Military HIV Natural History Study (NHS), a prospective cohort study, consisting of over 6,600 PWH in the military followed since the time of recruitment through HIV acquisition and during long-term ART suppression.^52^ Eligibility criteria were: (1) confirmed HIV-1 infection, (2) availability of plasma sample at least 1 year of ART suppression (i.e., plasma HIV RNA below the limit of assay detection at sample timepoint and at least one year after ART suppression), (3) availability of peripheral blood mononuclear cell (PBMC) sample to perform DNA extraction and whole genome sequencing.

For the Mendelian Randomization (MR) analysis, linking host genetics to vascular outcomes, we identified all eligible vascular event cases (N=205) and randomly selected cohort controls (N=788) from participants meeting inclusion criteria. Identifying a large number of vascular event cases remains a challenge even within the largest HIV cohorts. These cohorts have generally been established since the emergence of the HIV epidemic in the late 1980s and 1990s and often enrolled relatively young participants at study entry.^123–126^ Consequently, as in prior HIV studies,^127–129^ we analyzed incident vascular disease using composite outcomes to enhance statistical power. We defined atherosclerotic cardiovascular disease (ASCVD) as any new diagnosis of coronary artery disease (CAD), myocardial infarction (MI), cerebrovascular accident (CVA), or peripheral arterial disease (PAD). We also evaluated a broader composite outcome, vascular events (VE), which included ASCVD as well as venous thrombotic events – deep vein thrombosis (DVT) or pulmonary embolism (PE). We then performed participant selection using a case-cohort design. Given that both age and male sex are established risk factors for vascular disease,^130^ and since participants in the NHS cohort were relatively young and enriched for males, we oversampled controls to achieve comparable age and sex distributions. Eligible plasma samples were collected ≥1 year after achieving ART suppression. For cases, the qualifying plasma sample had to be collected at least one year prior to the incident vascular event to minimize potential reverse causation bias (e.g., inflammatory biomarker elevation following rather than preceding a vascular event). The total study population for our VE analyses were therefore comprised of 133 VE cases (including 110 ASCVD) and 694 controls (**Supplementary Fig. 10**; **Supplementary Table 15**).

### Plasma cytokine quantification

We quantified circulating plasma concentrations of IL-6, IL-1β, and IL-18 using the U-PLEX assay (Meso Scale MULTI-ARRAY Technology), which consists of SULFO-TAG™ conjugated antibodies and electrochemiluminescence detection. In all these assays, undiluted samples were run in duplicate following manufacturer’s instructions, and protein concentrations were determined using MSD Discovery Workbench (version 4.0.13) analysis software. The light intensities from the samples were interpolated using a four-parameter logistic fit (FourPL) to a standard curve of electrochemiluminescence generated from eight calibrators of known concentrations. The lower limit of detection for each marker can be found on the manufacturer’s website (https://www.mesoscale.com/∼/media/files/handout/assaylist.pdf). Batch correction was done on log-transformed cytokine values using the ComBat method from the R package sva.

### Whole genome sequencing and quality control analyses

Genomic DNA was isolated from cryopreserved peripheral blood mononuclear (PBMC) samples using standardized DNA isolation kits from Qiagen. DNA was prepared in aliquots with a normalized concentration and stored at -80°C. Whole genome sequencing using paired end reads with a target genome coverage of 60X was conducted on the Illumina NovaSeq system. We trimmed adapter sequences using *Trimmomatic* version 0.39, and we used the Burroughs-Wheeler Aligner (BWA) tool^131^ and the GenomeAnalysisToolkit (GATK) HaplotypeCaller joint variant calling method^132,133^ to perform DNA alignment. Reads were mapped to the human genome assembly GRCh38 using Picard tools.^134^ SNPs and insertions or deletions (indels) were then filtered by variant quality score recalibration (VQSR) using GATK.^135^ The whole genome data analysis toolset, PLINK,^136^ was then used to validate the chromosomal sex of each individual, filter out individuals with excessive heterozygosity, and SNPs violating Hardy-Weinberg equilibrium (HWE) at a p-value threshold of 1e-08. The VCFtools suite of functions were then used for data conversion and filtering out relevant SNPs.^137^

To stratify our cohort based on ancestry population, we referenced the 1000 Genomes Project,^138^ which gathered genotypes of 2,504 individuals from 26 different populations across the globe and classified 5 ancestry superpopulations: Europeans (EUR), Africans (AFR), South Asians (SAS), East Asians (EAS), and Admixed Americans (AMR).^139^ Individual genetics were mapped to the 1000 Genomes^138^ principal components and were assigned to the superpopulation of the nearest 1000 Genomes individual. We defined our cohort labels based on individuals mapped to the EUR and AFR superpopulations, given that these were the largest homogenous ancestry groups within our study (**Fig. 1B**; **Supplementary Fig. 1**). From each population, we removed individuals that were related (kinship estimate > 0.125). We imputed sex for each individual using the *plink --impute-sex* command and removed individuals with ambiguous sex (0.7 > F > 0.3). All subsequent analyses were performed stratified on these two cohorts, EUR and AFR. Within each cohort, we filtered out variants that violated Hardy-Weinberg Equilibrium (p<1e-06) and variants with a low call rate across individuals (>5% missingness). To control for population stratification, we performed principal component analysis (PCA) separately for the EUR and AFR cohorts, which were then included as covariates in all final models. To further control for genetic relatedness, we estimated the relatedness between each pair of participants within each EUR or AFR cohort using the *pcrelateToMatrix* function from *GENESIS.*^140^ We then included the genetic relationship matrix (GRM) within each of our mixed-effect association models for additional rigor in controlling for population stratification.

To evaluate potential residual population stratification, we examined the relationship between minor allele frequency (MAF) and SNP effect estimates, following an approach previously described by Sohail et al.^82^ For each cytokine (IL-6, IL-1β, and IL-18), SNP-level effect estimates from ancestry-specific association analyses were stratified by ancestry group (EUR and AFR). SNPs were grouped into 60 bins according to MAF, and the mean effect of SNPs on log-transformed cytokine levels was calculated within each bin. Only bins containing at least 300 SNPs were included to ensure stable estimates. Heat maps were generated to visualize MAF-dependent patterns in SNP effect sizes across cytokines and ancestry groups. Systematic diagonal banding was interpreted as evidence of residual population stratification, whereas the absence of such patterns indicated minimal confounding due to population structure. Consistent with this expectation, no evidence of residual stratification was observed (**Supplementary Fig. 12**).

### Single variant and gene-based association analyses

A total of 73,658,186 variants were sequenced from the original 1000 participants (**Supplementary Fig. 1**). After filtering for sequencing read depth and quality, a total of 66,084,459 variants from the 993 participants passed quality control for further analyses. After additional quality control filtering for potential admixture (PC1 < -0.112 for AFR and PC2 < -0.09 for EUR cohorts, respectively) and genetic relatedness (2 EUR and 1 AFR participant with kinship estimate >0.125), we were left with 417 participants in the EUR population and 387 participants in the AFR population. Final models demonstrated lambda genomic inflation factor (λ_GC_)^141^ values close to 1 (0.99 ± 0.0087 95% confidence interval), reflecting adequate adjustment for possible population stratification bias (**Supplementary Fig. 3**).

We performed individual variant analyses using the mixed effects modeling tool *GMMAT* (version 1.4.2).^70^ For each of the three cytokine phenotypes of interest (IL-6, IL-1β, and IL-18), associations were tested using imputed, batch-corrected cytokine measurements on the natural logarithmic scale. Because many participants contributed multiple cytokine measurements across different time points, we modeled the longitudinal structure of the data using a mixed-effects model with both a random intercept and a random slope for age at sample collection (random.slope = “AGE_AT_SAMPLE”). This approach accounts for within-individual correlation over time and allows for individual-specific temporal trends, such as transient inflammatory responses following clinical events. The European (EUR) cohort comprised 654 samples, and the African (AFR) cohort comprised 616 samples. All models were adjusted for age, sex, and the first five genetic principal components to account for population structure. For downstream functional annotation, we examined a ±500 kb window surrounding variants with association p < 1e-04 and queried the GTEx database to identify significant expression quantitative trait loci (eQTLs) across all available tissues.

To complement the single-variant analyses, we conducted gene-based rare variant association testing using the SMMAT framework implemented in GMMAT.^70^ Variants included in the gene-based tests comprised single-nucleotide variants and small insertions/deletions annotated as having HIGH predicted functional impact, as well as missense variants with a REVEL score > 0.6, based on Variant Effect Predictor (VEP) annotations. Variants failing Hardy–Weinberg equilibrium (p<1e-06) were excluded. Gene-based models incorporated the same covariates and genetic relationship matrix (GRM) used in the single-variant analyses, ensuring consistency across analytical approaches and appropriate control for relatedness and population structure. To reduce the likelihood of spurious associations driven by extremely sparse variant sets, analyses were restricted to genes containing at least four qualifying rare variants. Statistical significance was defined using a Bonferroni correction based on the number of genes tested for each phenotype (p < 1/number of genes analyzed).

### Transcriptome-wide association analyses

To investigate whether genetic associations with NLRP3-inflammasome–related cytokine levels are mediated through gene expression and to provide mechanistic insight into tissue-specific regulatory pathways underlying inflammasome-associated cytokine variation, we performed transcriptome-wide association analyses (TWAS) using our whole genome sequencing data.^142^ TWAS leverages genotype data to predict individual-level gene expression across tissues and subsequently tests the association between predicted expression and phenotypes of interest.

Predicted gene expression models were generated for four tissues relevant to immune and cardiovascular biology: coronary artery and left ventricular heart tissue, given their relevance to cardiovascular pathology; spleen, as a major immune organ; and whole blood, reflecting circulating inflammatory cytokine activity. Associations between predicted gene expression and plasma levels of IL-6, IL-1β, and IL-18 were evaluated separately in AFR and EUR ancestry cohorts.

Predicted gene expression was generated using GTEx v8 elastic net models implemented in *MetaXcan* (https://github.com/hakyimlab/MetaXcan). For association testing, we used the mixed-effects model framework from GMMAT, substituting predicted gene expression as the independent variable in place of genotype dosages. Analyses were restricted to four tissues selected for their relevance to cytokine biology and cardiovascular risk: whole blood, spleen, and heart left ventricle and coronary artery. We performed TWAS separately for EUR and AFR ancestry cohorts, for each of the three cytokines (IL-6, IL-1β, IL-18), and across the four tissue-specific expression models, yielding a total of 24 distinct TWAS analyses.

### Gene set enrichment analyses

For all analyses, we also conducted the gene set enrichment analysis (GSEA) to further summarize potential immune pathways associated with plasma cytokine levels. Genes were first rank-ordered by p-values. The rank-ordering was then used to identify immunologic pathways that were enriched from our dataset with *ClusterProfiler* using the MSigDB Hallmark, Canonical Pathways (CP), and Gene Ontology Biological Processes (GO-BP) databases.^143^ Pathways were filtered at a p-value threshold of 0.05 and visualized with network-based text-mining approach *vissE* (https://bioconductor.org/packages/vissE).

### Mendelian randomization analyses

To investigate the potential causal role of NLRP3-inflammasome–associated cytokines in vascular outcomes, we performed one-sample Mendelian randomization (MR) analyses^144^ stratified by ancestry. MR uses genetic variants as instrumental variables to estimate causal effects of an exposure (cytokine levels) on an outcome (vascular events), under established assumptions of relevance, independence, and exclusion restriction.^24,145^

Eligible incident vascular event cases (N = 205) and randomly selected controls (N = 795) were identified from the cohort. To increase statistical power given the limited number of events, we defined compositive outcomes. Atherosclerotic cardiovascular disease (ASCVD) included incident coronary artery disease (CAD), myocardial infarction (MI), cerebrovascular accident (CVA), or peripheral arterial disease (PAD), while a broader vascular events (VE) outcome additionally incorporated venous thrombotic events (VTE), including deep vein thrombosis (DVT) and pulmonary embolism (PE). After excluding individuals with events prior to within 12 months of plasma sampling, the final analytic set included 133 VE cases (110 ASCVD) and 694 controls (**Supplementary Fig. 10**; **Supplementary Table 15**). The final populations consisted of 337 total individuals with 64 VE (including 59 ASCVD) cases in the EUR population and 326 total individuals with 42 VE (including 32 ASCVD) in the AFR population. Ancestry-stratified analyses included 337 EUR participants (64 VE, 59 ASCVD) and 326 AFR participants (42 VE, 32 ASCVD).

We conducted one-sample MR analyses stratified by the EUR and AFR ancestry cohorts, as previously described.^146,147^ ASCVD was the primary outcome (given the established role of the NLRP3 inflammasome in atherogenesis) and VE (which includes VTE) was analyzed as the secondary outcome. Causal effects were estimated using the inverse-variance weighted (IVW) method as the primary approach, and MR-Egger, weighted median, and MR-PRESSO methods were applied as complementary sensitivity analyses to assess robustness to violations of instrumental variable assumptions The IVW method provided the most efficient estimates under the assumption that horizontal pleiotropy was absent or balanced across variants. MR-Egger regression allowed for directional (unbalanced) pleiotropy under the assumption that the strength of the association between variants and the exposure was independent of pleiotropic effects. The weighted median and MR-PRESSO approaches provided valid estimates when at least 50% of the variants satisfied instrumental variable assumptions.

Genetic instruments for each cytokine were selected based on association with plasma levels (p<1e-05), minor allele frequency (MAF>0.01), and instrument strength (F-statistic>10), and were pruned for linkage disequilibrium (250 kb window, r²<0.5). Variant-exposure and variant-outcome associations were estimated using GMMAT, adjusting for age, sex, and the first five principal components. Traditional cardiovascular (hypertension, hyperlipidemia, type II diabetes, family history, tobacco/alcohol use) and HIV-related covariates (nadir CD4⁺ T-cell count, log₁₀ pre-ART viral load, ART timing, time to viral suppression, duration of suppression) were evaluated but not included in the final models, as our analyses suggested that these variables may lie along the causal pathway from genetic variation to vascular outcomes and could therefore violate core MR assumptions.^148^

To evaluate potential violations of MR assumptions, we assessed heterogeneity using Cochran’s Q and horizontal pleiotropy using MR-Egger intercepts and MR-PRESSO. No significant heterogeneity (differences in SNP-specific effects of the cytokine on vascular events) was observed across cytokine-outcome combinations (**Supplementary Table 17**). MR-Egger intercepts showed minimal evidence of directional pleiotropy overall, with the exception of IL-1β in AFR participants (**Supplementary Table 18**). Sensitivity analyses using MR-PRESSO detected no influential outliers in the IL-1β AFR analysis, and effect estimates were consistent across methods. Taken together, these findings indicated that the genetic instruments were robust and that MR assumptions were largely satisfied. Accordingly, IVW estimates were used as the primary measure of causal effect, with complementary methods providing consistent support for the observed associations.^149,150^

### CRISPR-based functional validation

To annotate genome-wide loci identified in our study, we integrated gene sets derived from single-variant (GWAS; p<1e-05), rare variant, and TWAS analyses with two publicly available CRISPR-based functional datasets generated in primary human CD4⁺ T cells.

First, we used gene-level statistics from a genome-wide CRISPRa/CRISPRn screen that systematically identified pro- and anti-HIV host factors in primary human CD4⁺ T cells.^151^ In this study, transcriptional activation (CRISPRa) of >18,800 genes and knockout (CRISPRn) of >19,100 genes were performed in primary CD4⁺ T cells, followed by infection with replication-competent GFP-tagged HIV (NLENG1I, an NL4-3 derivative). Cells were sorted by FACS into HIV-GFP⁺ (infected) and HIV-GFP⁻ (uninfected) populations. Gene-level log₂ fold-changes (LFC) and FDR were computed using MAGeCK v0.5.9.5 based on sgRNA count differences between these populations. In the CRISPRa screen, positive LFC indicated enrichment in infected cells (pro-viral effect), whereas negative LFC indicated enrichment in uninfected cells (anti-viral effect); the directional interpretation was reversed for CRISPRn. Genes were classified as pro-viral, anti-viral, or non-hits based on FDR<0.2 (Table S2 of the original Zhu et al.^151^ study).

Second, we cross-referenced our hits against a genome-scale Perturb-seq dataset in primary human CD4⁺ T cells.^90^ In that study, naive CD4⁺ T cells from four donors were transduced with a CRISPRi library targeting all expressed genes, and transcriptomes were profiled across approximately 22 million cells in three conditions: rest (Rest), 8 hours after re-stimulation (Stim8hr), and 48 hours after re-stimulation (Stim48hr), using a probe-based 10x Flex single-cell RNA-seq platform. Differential expression effects of each perturbation on each measured gene were estimated as log_2_ fold-changes (log2FC) of perturbed versus non-targeting control pseudobulks, with associated p-values and Benjamini-Hochberg FDR.

We performed two complementary analyses using the perturbation × gene expression matrix (Fig. 2C of Zhu et al.^151^). In the upstream analysis, we tested whether perturbation of candidate genes altered expression of *IL6*, *IL1R1*, or *IL18R1* across stimulation conditions, using these transcripts as proxies for cytokine production or receptor availability. Because *IL1B* and *IL18* transcript measurements were not available in this dataset, *IL1R1* and *IL18R1* were used as surrogate readouts, which represents a limitation. In the downstream analysis, we evaluated whether perturbation of *IL6R*, *IL1B*, or *IL18R1* altered expression of candidate genes, to assess reciprocal regulatory relationships.

For both datasets, we quantified overlap between study-derived gene sets and CRISPR-identified hits using Fisher’s exact test (one-sided, alternative = “greater”) and an empirical permutation framework. The background set was restricted to genes tested in both the WGS analyses and the respective CRISPR dataset. For each comparison, 10,000 random gene sets matched in size to the input gene list were sampled from the background, and empirical p-values were calculated as the proportion of permutations with overlap equal to or greater than observed. Analyses were performed across cytokines, ancestry groups, and gene sets (GWAS, rare variant, TWAS, and combined), and Benjamini–Hochberg correction was applied within each dataset.

CRISPRa/CRISPRn hits were defined as FDR<0.2 based on MAGeCK output. For the Perturb-seq dataset, where no candidate genes met this threshold for cytokine targets, perturbation effects were evaluated at nominal p<0.05 for upstream analyses and p<0.05 with |log₂FC|>0.2 for downstream analyses to incorporate effect size filtering. Individual gene-level effects were summarized descriptively given differences in experimental context, including cell state, perturbation modality, and measured phenotypes.

### Validation of GWAS findings of IL-6 levels on an external cohort

To assess the generalizability of IL-6 GWAS findings, we performed validation analyses in participants from the All of Us Research Program (Controlled Tier Dataset v8)^99^. Analyses were restricted to individuals without HIV infection or malignancy and with available plasma IL-6 measurements and genome-wide genetic data. IL-6 concentrations were log-transformed prior to analysis. After phenotype cleaning and merging with genetic data, 570 participants from All of Us were retained for analysis.

Whole-genome sequencing data (GRCh38) from All of Us were processed using Hail. Variants were restricted to biallelic SNPs with call rate ≥95%, and linkage disequilibrium pruning was performed prior to principal component analysis in PLINK2. Genetic ancestry was assigned using the All of Us ancestry prediction model,^99^ and analyses were stratified by European (EUR; N = 241) and African (AFR; N = 83) ancestry groups to minimize confounding due to population structure.

Genome-wide association testing was performed using linear score tests implemented in GMMAT on log-transformed IL-6 levels, adjusting for age, sex, and ancestry principal components. Models including 0-5 principal components were evaluated based on genomic inflation (λ_GC_) ^152^ (**Supplementary Table 23**), and final models included PC1-PC2 for EUR participants and PC1 for AFR participants. Variants passing analysis-level quality control included 305,827 variants in EUR and 437,855 variants in AFR. Genomic inflation factors for the final models were λ_GC_= 1.0075 (EUR) and λ_GC_ = 1.0253 (AFR) indicating adequate control for population structure.

Linear mixed models incorporating a genomic relatedness matrix (GRM) were additionally evaluated to account for cryptic relatedness. Because inclusion of the GRM did not materially improve genomic control and resulted in unstable convergence in EUR analyses, primary results are reported from fixed-effect models without kinship correction.

Gene-level association statistics were derived by mapping each SNP to its nearest protein-coding gene using pyranges (22,250 protein-coding genes; no fixed distance cutoff). Preranked GSEA was performed against curated KEGG pathway gene sets using gseapy^153,154^, retaining pathways with 15–500 genes present in the ranked gene list (292 and 291 pathways in EUR and AFR, respectively). Pathway enrichment results are reported at an exploratory FDR threshold of 0.25.

Cross-cohort pathway concordance was assessed by Spearman rank correlation of normalized enrichment scores. Pathway overlap was evaluated using a gene-level permutation framework (10,000 permutations) in which discovery cohort gene-level ranking statistics were randomly reassigned to genes without replacement, preranked GSEA was rerun on each permuted list, and overlap with observed validation cohort significant pathways was recorded. Two-tailed empirical P-values were calculated as 2 x *min*(*P*(*null* > *observed*), *P*(*null* < *observed*)), capped at 1.0.

Cross-cohort SNP-level comparisons were performed after harmonizing variants by genomic position and allele annotation, excluding variants with non-matching alleles. An asymmetric threshold framework was applied, evaluating variants suggestive in the discovery cohort (P < 1e-05) against nominal significance in the validation cohort (P < 0.05). All genotype processing used Hail v0.2.134 and PLINK2; association testing used GMMAT in R; and gene- and pathway-level analyses used gseapy, pyranges, and standard Python libraries.

## Data availability

Data underlying the findings reported in this article are available from the corresponding author upon reasonable request, subject to institutional review and approval and completion of a data use agreement. Deidentified participant data, along with a data dictionary, will be made available to qualified investigators for purposes of replicating procedures and results.

## Code availability

The code to reproduce our analyses and figures is available through the Zenodo database (https://doi.org/10.5281/zenodo.20548783).

## ACKNOWLEDGEMENTS

The authors wish to acknowledge the participation of all the study participants who contributed to this work as well as the clinical research staff of the U.S. Military HIV Natural History Study cohort who made this research possible. They also gratefully acknowledge the contribution of Priscilla Y. Hsue related to the interpretation of HIV cardiovascular associations, and of David A. Siegel for the application of Generalized linear Mixed Model Association (GMMAT) analyses. This work was supported in part by the National Institutes of Health: K23GM112526 (SAL), NIH/NIAID R01AI143464 (SAL), NIH/NHGRI U24HG007346 (NI), UM1TR004409 (CTM), and NIH/NIAID U54AI170792 (AM). NI is a Chan Zuckerberg Biohub San Francisco Investigator. Partial support for this work (IDCRP-000-03) was provided by the Infectious Disease Clinical Research Program (IDCRP), a Department of Defense program executed through the Uniformed Services University of the Health Sciences, Department of Preventive Medicine and Biostatistics through a cooperative agreement with The Henry M. Jackson Foundation for the Advancement of Military Medicine, Inc. (HJF). This project has been funded in part by the National Institute of Allergy and Infectious Diseases, National Institutes of Health, under Inter-Agency Agreement Y1-AI-5072, and the Defense Health Program, U.S. DoW, under award HU0001190002. The views expressed are those of the authors and do not reflect the official views of the Uniformed Services University of the Health Sciences, the Henry M. Jackson Foundation for the Advancement of Military Medicine, Inc., the National Institutes of Health or the Department of Health and Human Services, the Department of Defense, Defense Health Agency, the Departments of the Army, Navy or Air Force, and U.S. Government. The funders had no role in study design, data collection and analysis, decision to publish, or preparation of the manuscript.

## AUTHOR CONTRIBUTIONS STATEMENT

All authors provided critical feedback in finalizing the report. SAL conceived and designed the study. BKA obtained funding to support the clinical enrollment of study participants, and SAL obtained funding for the characterization of the host genetic and plasma cytokine data. BKA and XC coordinated the collection, management, and quality control processes for the clinical data, and BKA, ST, AG, JMY, DTL, TL, ECE, and REC provided the biospecimens. CTM and DW performed the PBMC processing and DNA extraction for whole genome sequencing. JAT performed the plasma sample processing and cytokine assays under the guidance of RPS. SAL, SSav, CS, SSar, MSD, and VP coordinated the management, adjudication, and quality control processes for accurately linking the clinical, cytokine, and genetic data for downstream analyses. SAL and ASB analyzed the clinical data with data quality control support from BKA, XC, SSav, CS, SSar, MSD, and VP. RC developed the GWAS and TWAS models, under the guidance of SAL, NI, and ASB. NSC performed the GSEA, and MR analysis under the guidance of SAL, ASB, and EL. CRISPR validation was performed by NSC under the guidance of ED, UR, AM, and SAL. SSho performed validation analysis using external public genomic database, under the guidance of SAL, EL, and NSC. RC, NSC, ASB, SSav, SSho, EL, and SAL finalized data visualization for the manuscript. SAL, RC, NSC, SSav, SSho, and EL wrote the report with critical feedback from the additional authors.

Correspondence should be addressed to SAL.

