## Supplemental Figures for "Host Genetic Regulation of NLRP3 Inflammasome Cytokines Reveals Immune and Vascular Pathways in HIV"

**Supplementary Fig. 1. Study participant selection and genetic ancestry.** After filtering for sequencing read depth and quality, a total of 66,084,459 variants from 993 participants passed quality control and were included in downstream analyses. Our study population largely included participants of European (EUR) and African (AFR) ancestry.

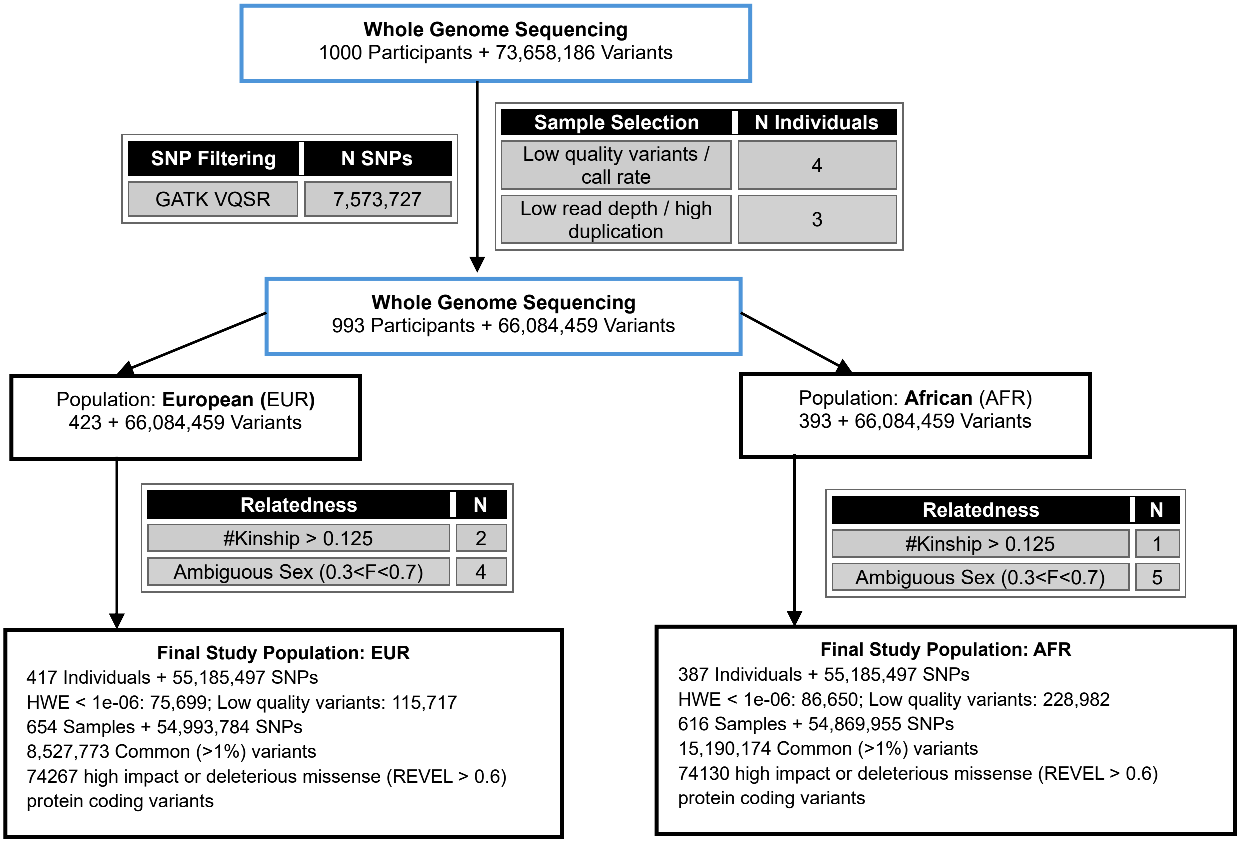

**Supplementary Fig. 2. Detection and correction of batch effects in plate standards.** Cytokine standards of known concentration were run on each of the 29 assay plates. Plates exhibiting batch effects – defined as having more than two standard measurements exceeding two median absolute deviations from the median standard value – are shown in black ("Outlier Plates"), whereas plates without evidence of batch effects are shown in green ("Typical Plates"). These results indicate the presence of plate-to-plate variability and highlight the need for batch correction to enable reliable comparisons of cytokine measurements across plates (**A**). These results suggest the need for batch-correction to ensure that cytokine measurements could be compared across plates in subsequent analyses. Observed cytokine concentrations by plate before (**B**) and after (**C**) batch correction are shown below. Density plots show the distribution of measurements from outlier (black) and typical (green) plates for each cytokine. After batch correction, the distributions from outlier and typical plates are more closely aligned, indicating successful mitigation of batch effects.

**A.**

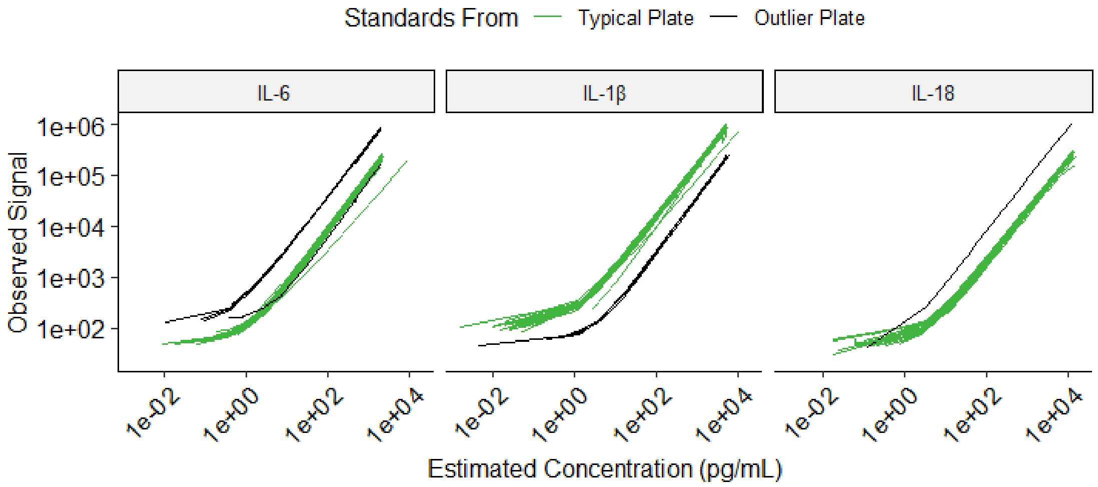


**B.**

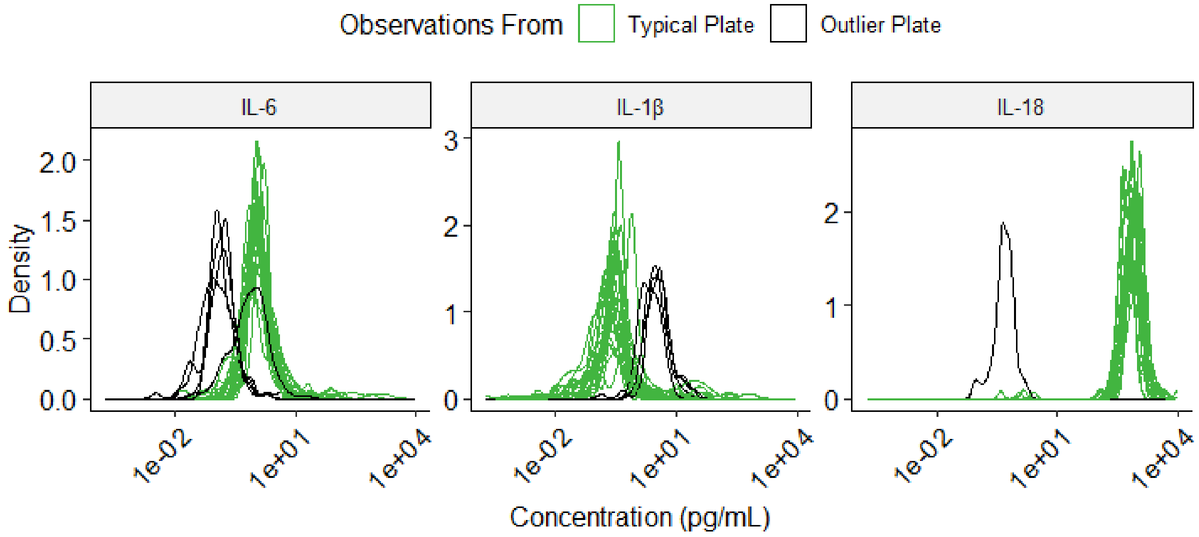


**C.**

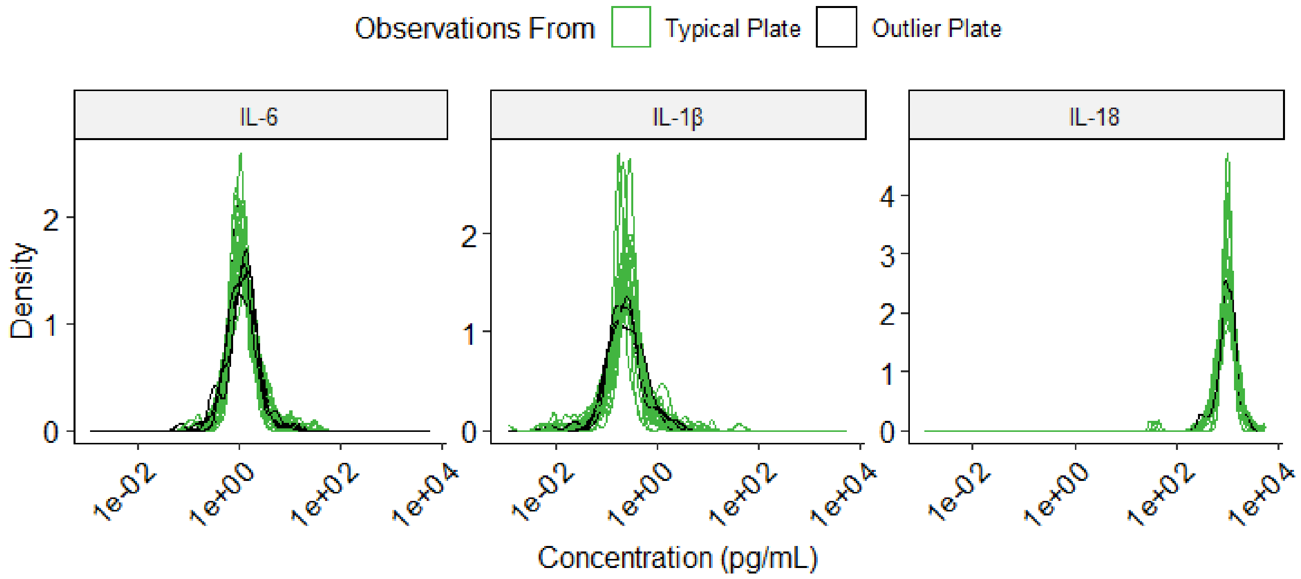

Observations From
Typical Plate
Outlier Plate
IL-6
IL-1β
IL-18
Density
0
1
2
3
4
1e-02
1e+00
1e+02
1e+04
Concentration (pg/mL)

**Supplementary Fig. 3. Quantile-quantile (Q-Q) plots from genome-wide association studies (GWAS) single variant analysis of NLRP3-inflammasome-associated cytokines: IL-6 (A), IL-1β (B), and IL-18 (C).** For each cytokine, results from the European (EUR) ancestry population are shown in the left panels, and results from the African (AFR) ancestry population are shown in the right panels.

**EUR** **AFR**

**A.**

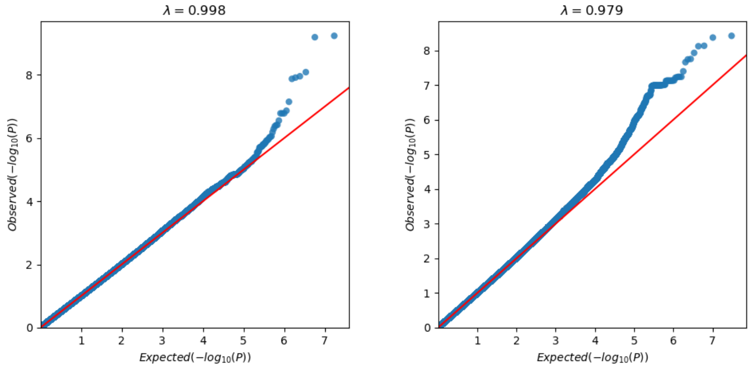


**B.**

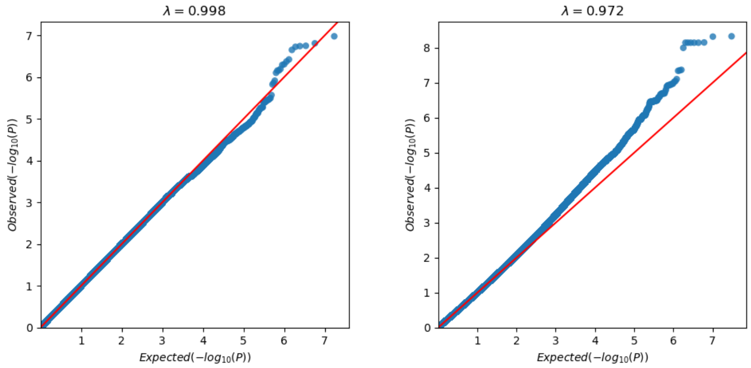


**C.**

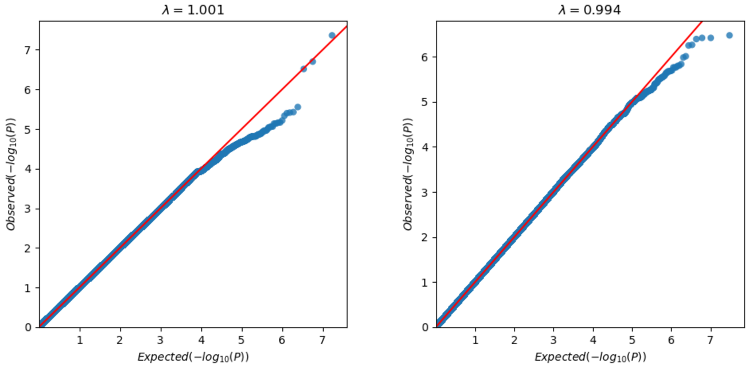

**Supplementary Fig. 4. Regional (locus) plots for each genome-wide association signal for plasma IL-6 in the European (EUR) ancestry population**. The lead (most significant) variant is annotated with its chromosome, position, reference allele, and alternative allele. The dotted horizontal line denotes the genome-wide significance threshold (p < 5e-08). Where available, variants are colored according to linkage disequilibrium (LD) with the lead variant, based on the 1000 Genomes EUR reference panel. Loci are named according to the nearest gene: *GOLGAL8S* (**A**) and *KLHL29* (**B**).

**A.**

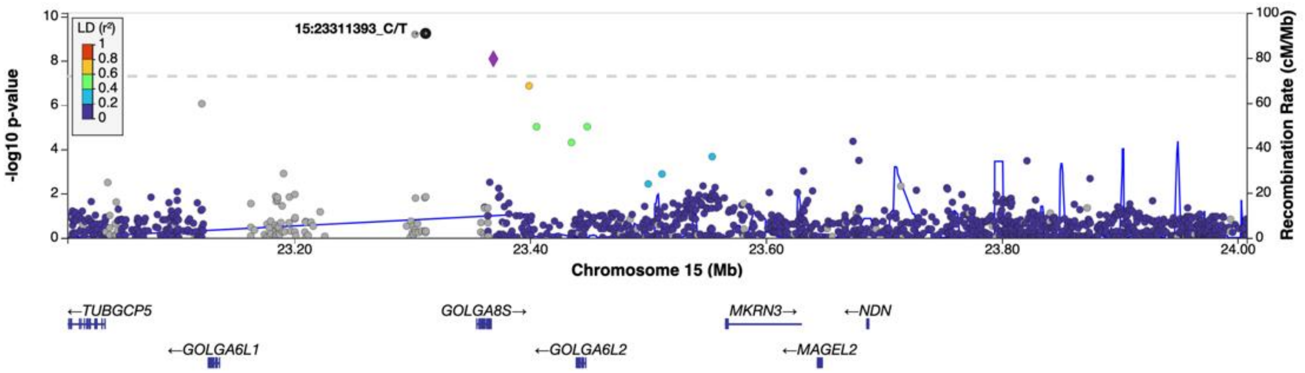


**B.**

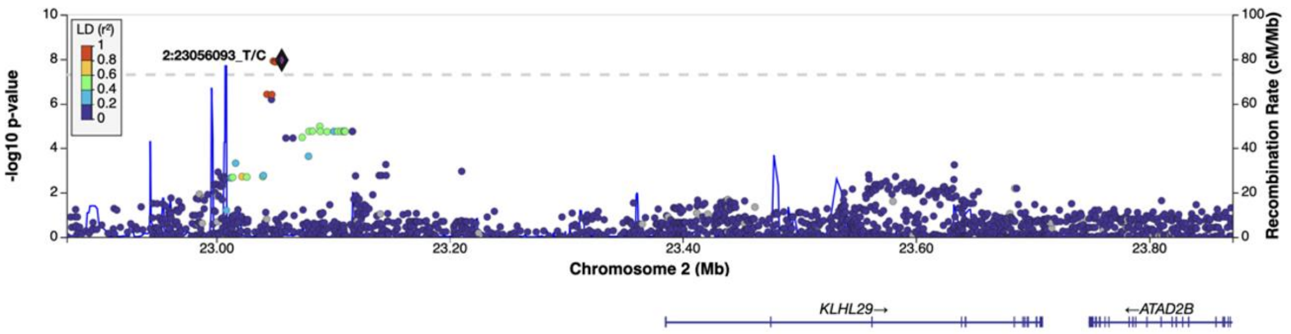

**Supplementary Fig. 5. Regional (locus) plots for each genome-wide association signal for plasma IL-6 in the African (AFR) ancestry population.** The lead (most significant) variant is annotated with its chromosome, position, reference allele, and alternative allele. The dotted horizontal line denotes the genome-wide significance threshold ($p < 5e-08$). Where available, variants are colored according to linkage disequilibrium (LD) with the lead variant, based on the 1000 Genomes AFR reference panel. Loci are named according to the nearest gene: *EPCAM* (**A**), *AGMO* (**B**), *EEA1* (**C**), *FGF9* (**D**), and *FTO/IRX3* (**E**).

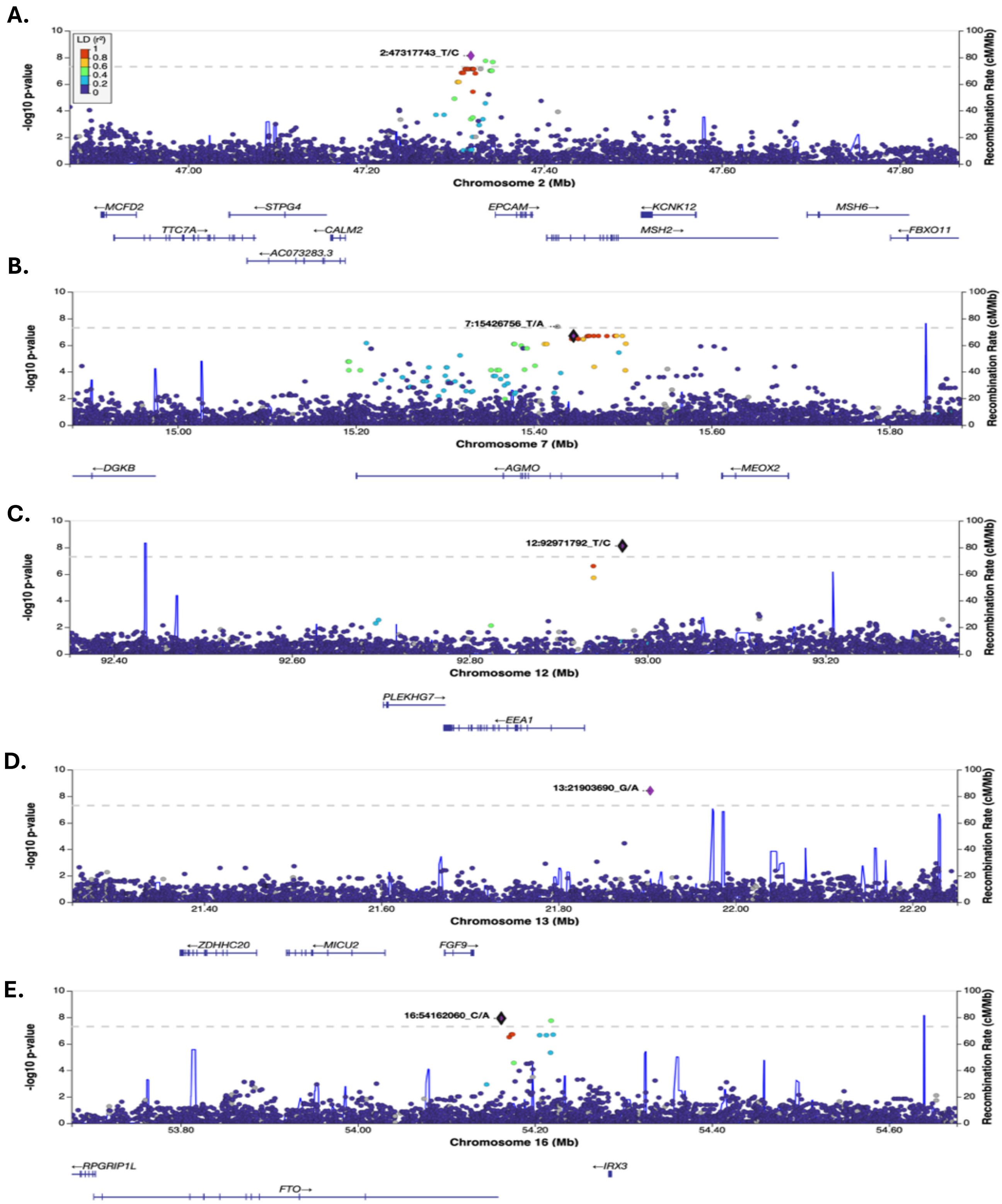

**Supplementary Fig. 6. Regional (locus) plots for each genome-wide association signal for plasma IL-1β in the African (AFR) ancestry population.** The lead (most significant) variant is annotated with its chromosome, position, reference allele, and alternative allele. The dotted horizontal line denotes the genome-wide significance threshold (p < 5e-08). Where available, variants are colored according to linkage disequilibrium (LD) with the lead variant, based on the 1000 Genomes AFR reference panel. Loci are named according to the nearest gene: *IGK* (**A**), *DGKB* (**B**)*, RALYL* (**C**)*, and MCTP2* (**D**).

**A.**

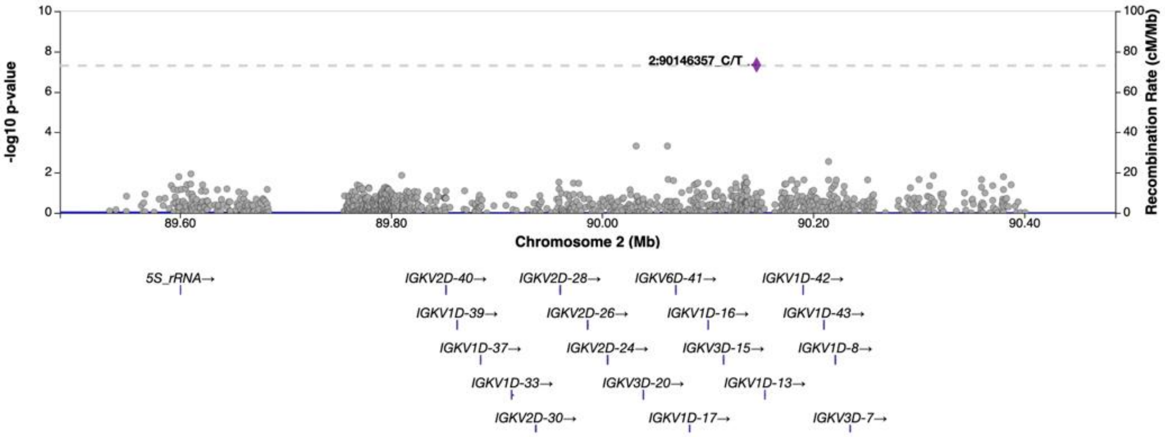


**B.**

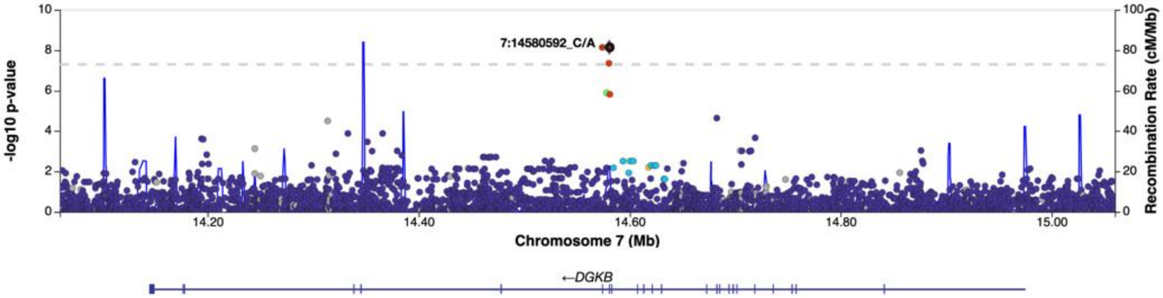


**C.**

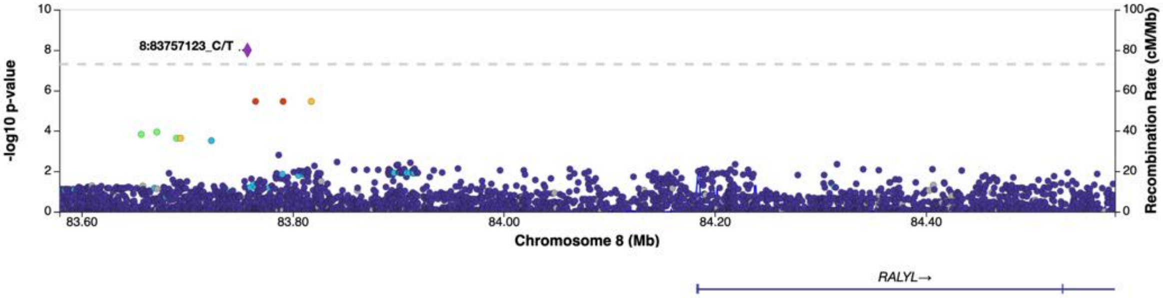


**D.**

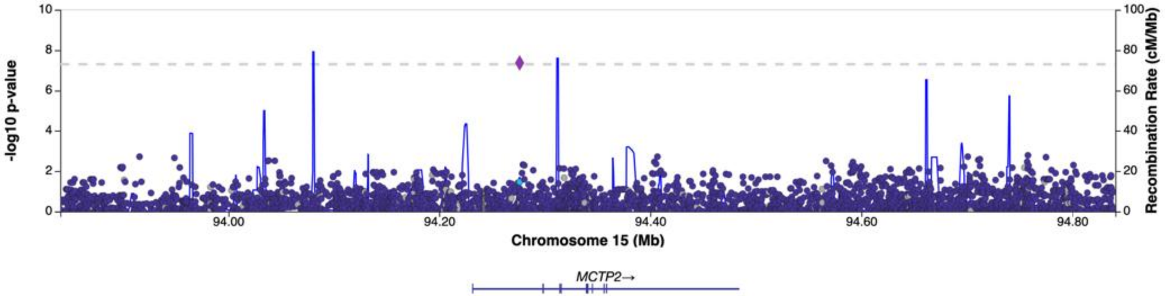

**Supplementary Fig. 7. Regional (locus) plots for each genome-wide association signal for plasma IL-18 in the European (EUR) ancestry population**. The lead (most significant) variant is annotated with its chromosome, position, reference allele, and alternative allele. The dotted horizontal line denotes the genome-wide significance threshold (p < 5e-08). Where available, variants are colored according to linkage disequilibrium (LD) with the lead variant, based on the 1000 Genomes EUR reference panel. Loci are named according to the nearest gene: *ETV1*.

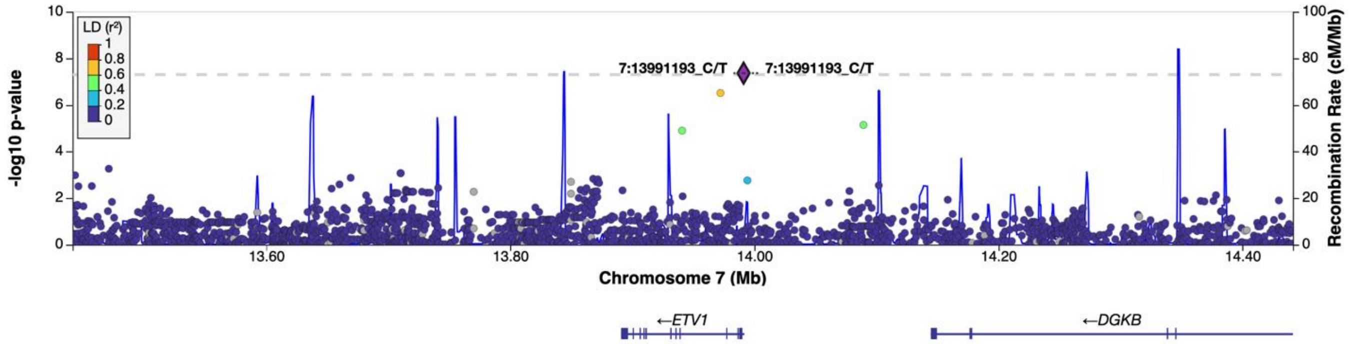

**Supplementary Fig. 8. Quantile-quantile (Q-Q) plots from gene-based rare variant analysis of NLRP3-inflammasome-associated cytokines: IL-6 (A), IL-1β (B), and IL-18 (C).** For each cytokine, results from the European (EUR) ancestry population are shown in the left panels, and results from the African (AFR) ancestry population are shown in the right panels.

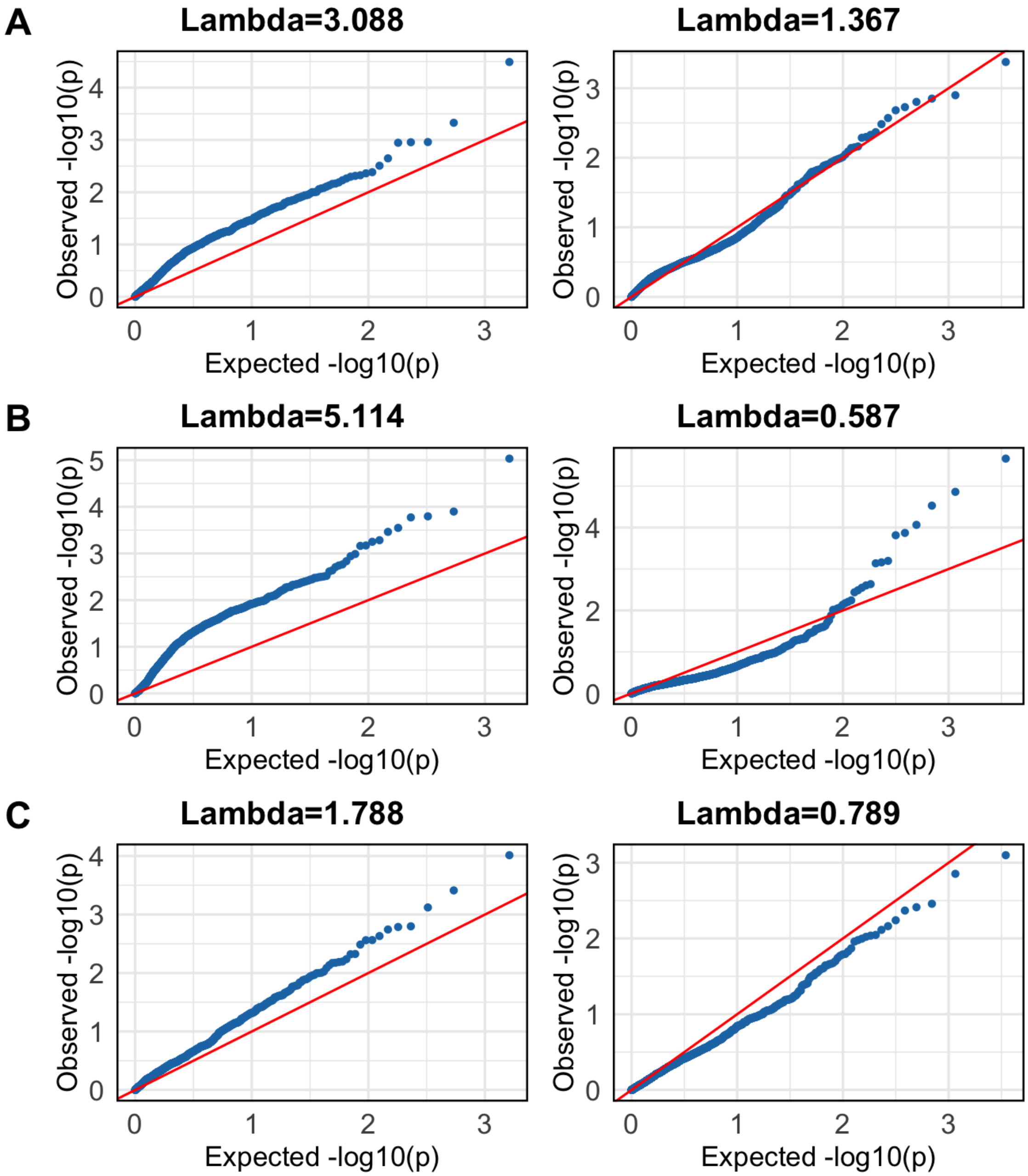

**Supplementary Fig. 9. Quantile-quantile (Q-Q) plots from transcriptome-wide association studies (TWAS) of NLRP3-inflammasome-associated cytokines: IL-6 (A), IL-1β (B), and IL-18 (C).** For each cytokine, results are shown for the European (EUR; top rows) and African (AFR; bottom rows) ancestry populations across four tissues: coronary artery (first column), left ventricular heart tissue (second column), spleen (third column), and whole blood (fourth column).

**A.**

**IL-6 (EUR)**

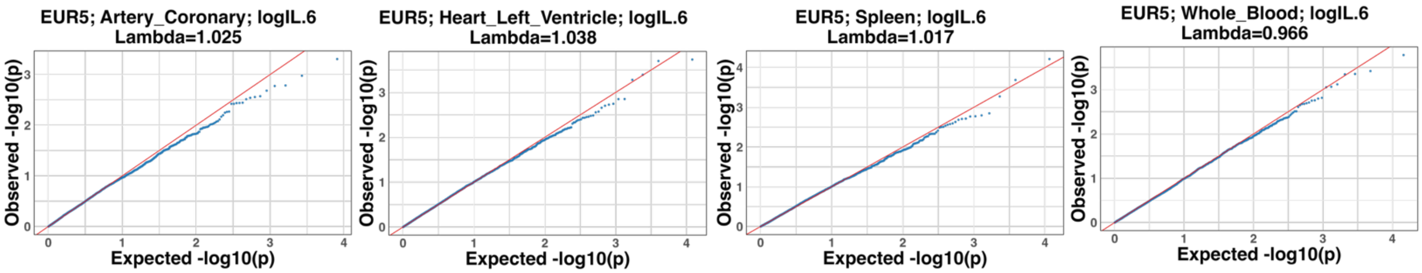


**IL-6 (AFR)**

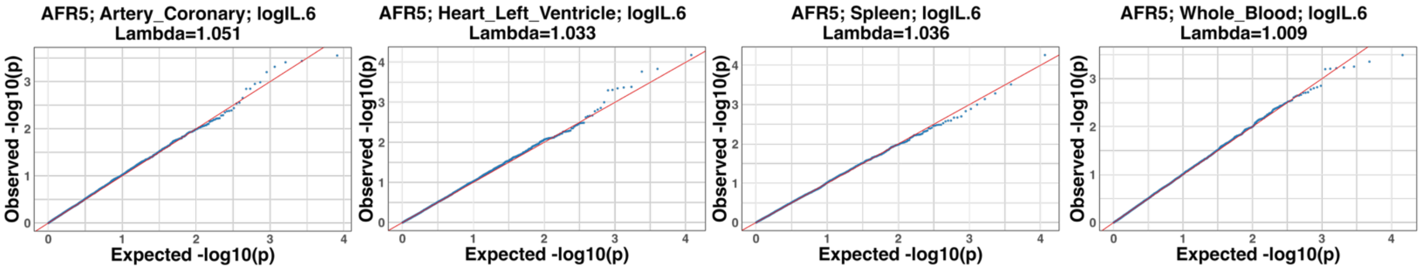


**B.**

**IL-1β (EUR)**

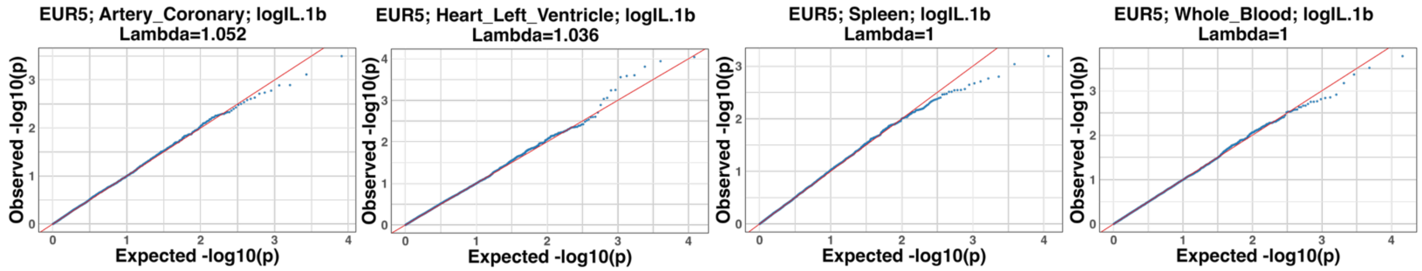


**IL-1β (AFR)**

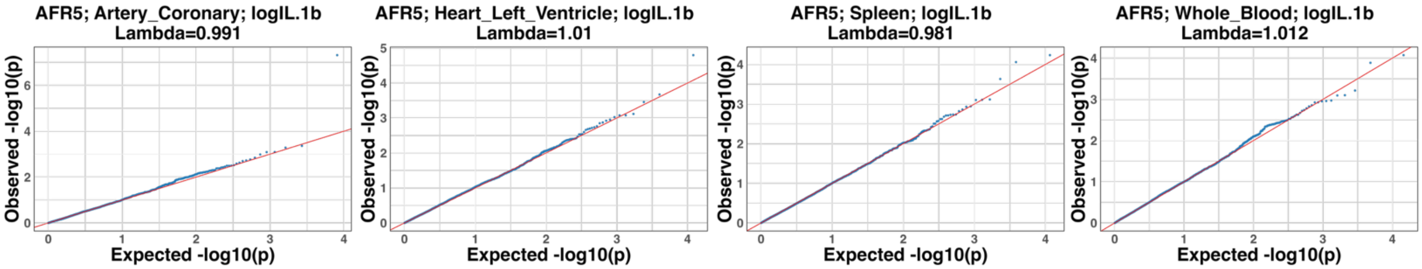

C.

### IL-18 (EUR)

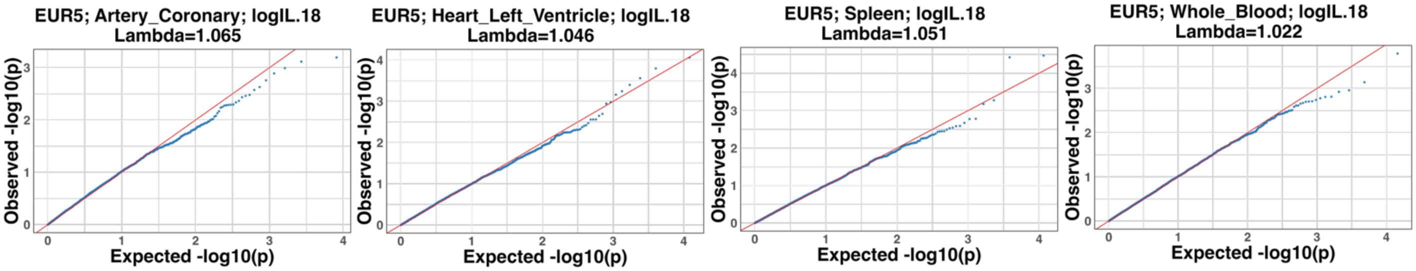


### IL-18 (AFR)

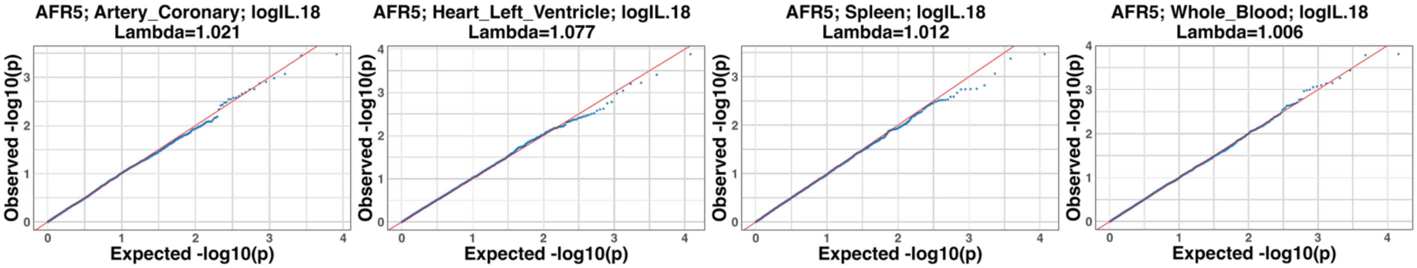

**Supplementary Fig. 10. Age, sex, race/ethnicity, and follow-up time of final selected case-cohort samples.** After quality control filtering for sequencing depth and variant call reliability, genetic data from 827 participants met criteria for inclusion in downstream analyses (see **Fig. 1**) and had at least 12 months between viral suppression and a plasma sample and between the plasma sample and either the first vascular event or censorship date. We used a stratified case-cohort design to select 1,002 participants. Since age and male sex are established risk factors for vascular disease, we oversampled controls to achieve comparable age and sex distributions across groups. Results of sample selection show relatively comparable age (at plasma sample) (**A**), follow-up time (**B**), sex (**C**), and race/ethnicity (**D**) distributions by group. ASCVD = atherosclerotic cardiovascular disease; any new onset coronary artery disease (CAD), myocardial infarction (MI), cerebrovascular accident (CVA), or peripheral arterial disease (PAD) diagnosis. Venous thrombotic events (VTE); any new onset deep vein thrombosis (DVT) or pulmonary embolism (PE). VE = vascular event; composite outcome of any incident ASCVD or VTE.

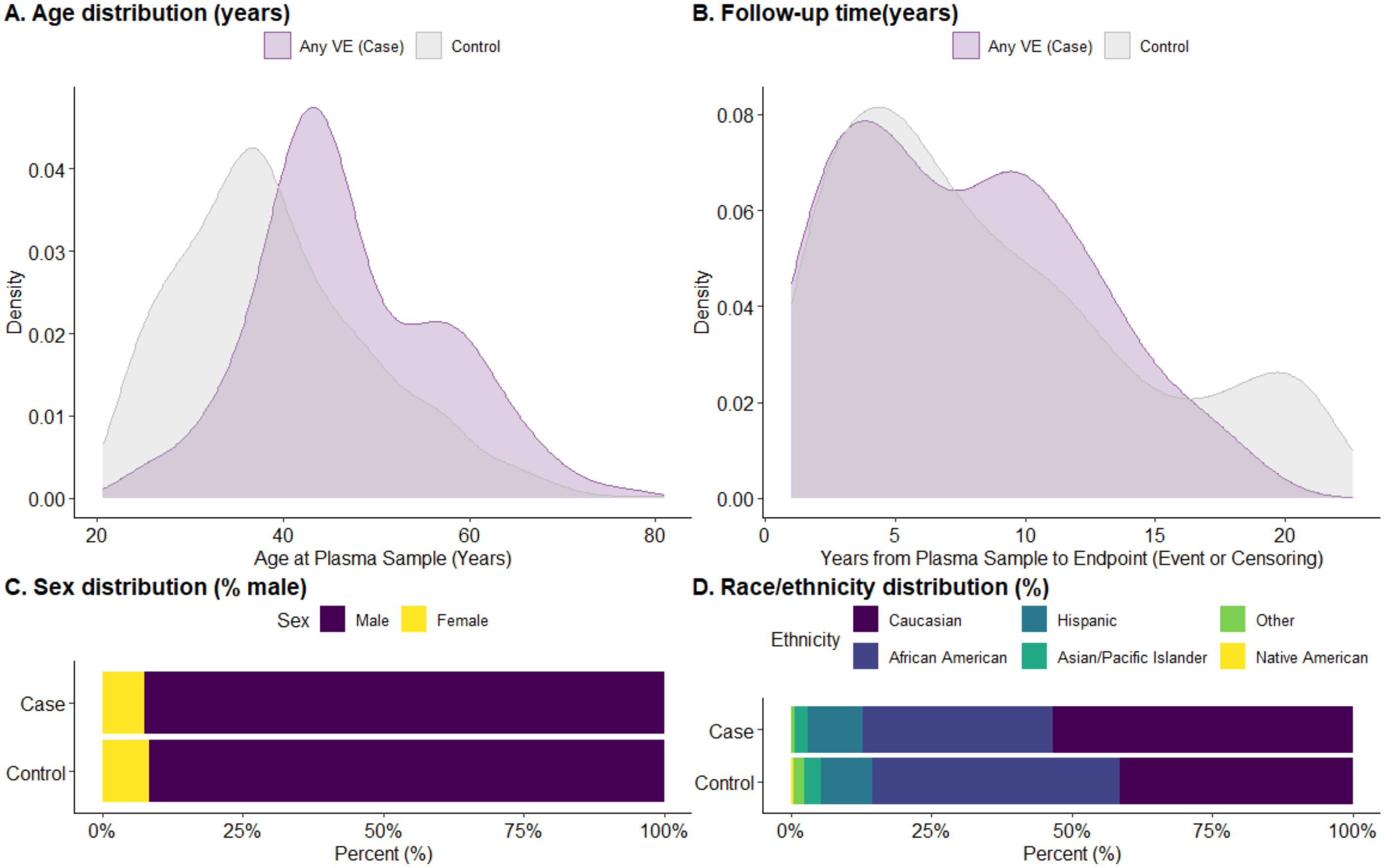

**Supplementary Fig. 11. NLRP3 inflammasome-associated cytokine concentrations across incident vascular diagnoses.** Disaggregated Mann-Whitney tests comparing plasma concentrations of IL-6 (**A**), IL-18 (**B**), and IL-1β (**C**) are shown for specific incident vascular event (VE) diagnoses. IL-6 and IL-18 levels were increased among participants with incident atherosclerotic cardiovascular disease (ASCVD; red boxes), with IL-6 and IL-18 significantly elevated among participants with CAD compared with controls. IL-6 and IL-18 were also somewhat elevated in patients with PAD, as well as IL-18 with CVA. In contrast, none of the cytokines differed significantly for either venous thrombotic event (VTE) outcome (blue boxes). ASCVD is defined as a composite of new-onset coronary artery disease (CAD), myocardial infarction (MI), cerebrovascular accident (CVA), or peripheral arterial disease (PAD). VE represents a broader composite outcome encompassing ASCVD as well as VTE, including deep vein thrombosis (DVT) and pulmonary embolism (PE).

**A. IL-6**

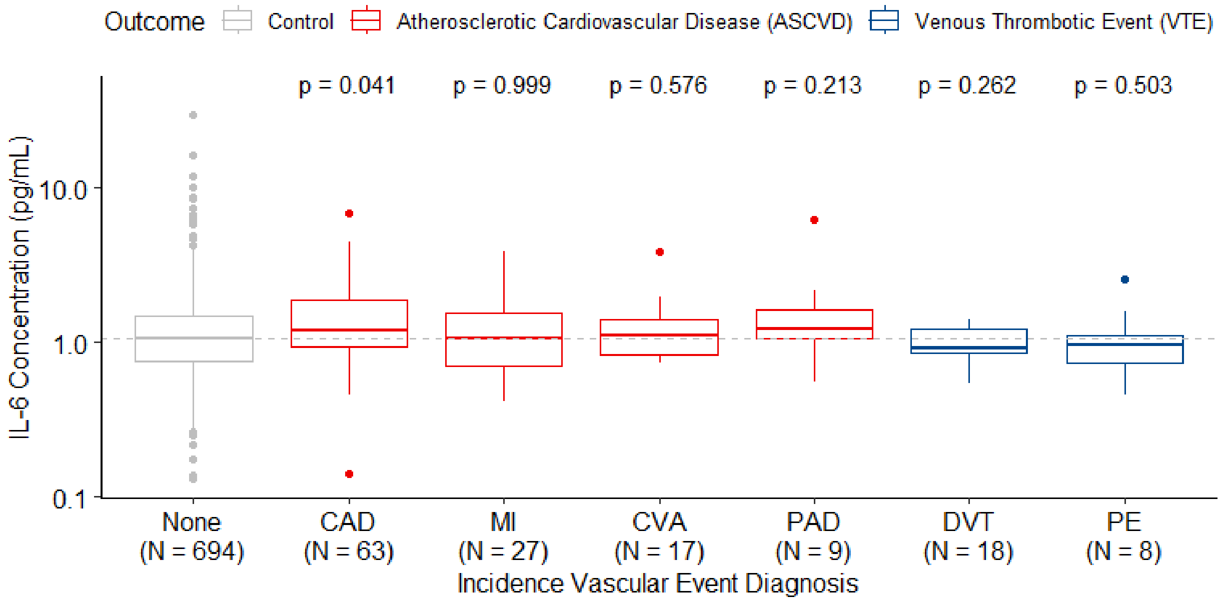


**B. IL-1β**

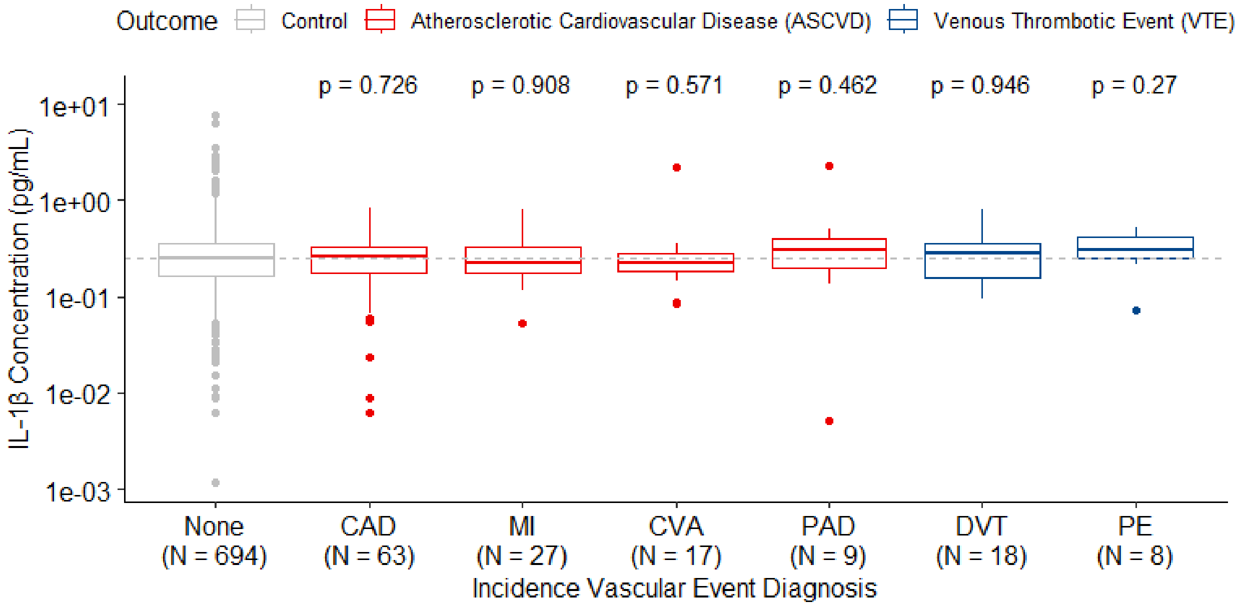
Outcome
Control
Atherosclerotic Cardiovascular Disease (ASCVD)
Venous Thrombotic Event (VTE)
p = 0.726
p = 0.908
p = 0.571
p = 0.462
p = 0.946
p = 0.27
1e+01
1e+00
1e-01
1e-02
1e-03
IL-1β Concentration (pg/mL)
None (N = 694)
CAD (N = 63)
MI (N = 27)
CVA (N = 17)
PAD (N = 9)
DVT (N = 18)
PE (N = 8)
Incidence Vascular Event Diagnosis

**C. IL-18**

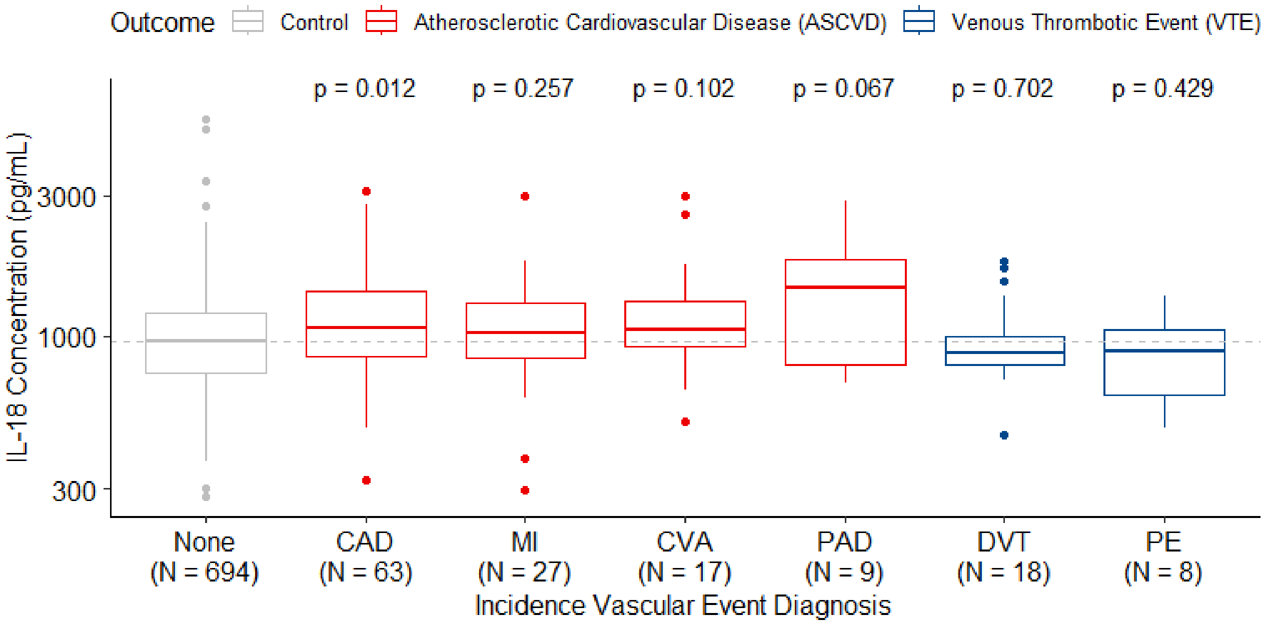
Outcome
Control
Atherosclerotic Cardiovascular Disease (ASCVD)
Venous Thrombotic Event (VTE)
p = 0.012
p = 0.257
p = 0.102
p = 0.067
p = 0.702
p = 0.429
3000
1000
300
IL-18 Concentration (pg/mL)
None (N = 694)
CAD (N = 63)
MI (N = 27)
CVA (N = 17)
PAD (N = 9)
DVT (N = 18)
PE (N = 8)
Incidence Vascular Event Diagnosis

**Supplementary Fig. 12. Assessment of residual population stratification using allele frequency-stratified SNP effects.** Heat maps display the mean estimated effects of single-nucleotide polymorphisms (SNPs) on plasma IL-6 (**A**), IL-1β (**B**), and IL-18 (**C**) levels, shown separately for European (EUR) and African (AFR) ancestry groups. SNPs were binned into 61 groups according to minor allele frequency (MAF), and the mean SNP effect on log-transformed cytokine levels was calculated within each bin. Only MAF bins containing at least 300 SNPs are shown. This analysis was performed to evaluate potential residual population stratification, as previously described. If there is incomplete control of population structure, SNPs that are more common in one population than the other may be estimated to be more positively (or negatively) associated with the cytokine level in that population than the true effect, which would be displayed as positive values in the upper left diagonal half and negative values in the bottom right (or vice versa) of the plot.

**A. IL-6**

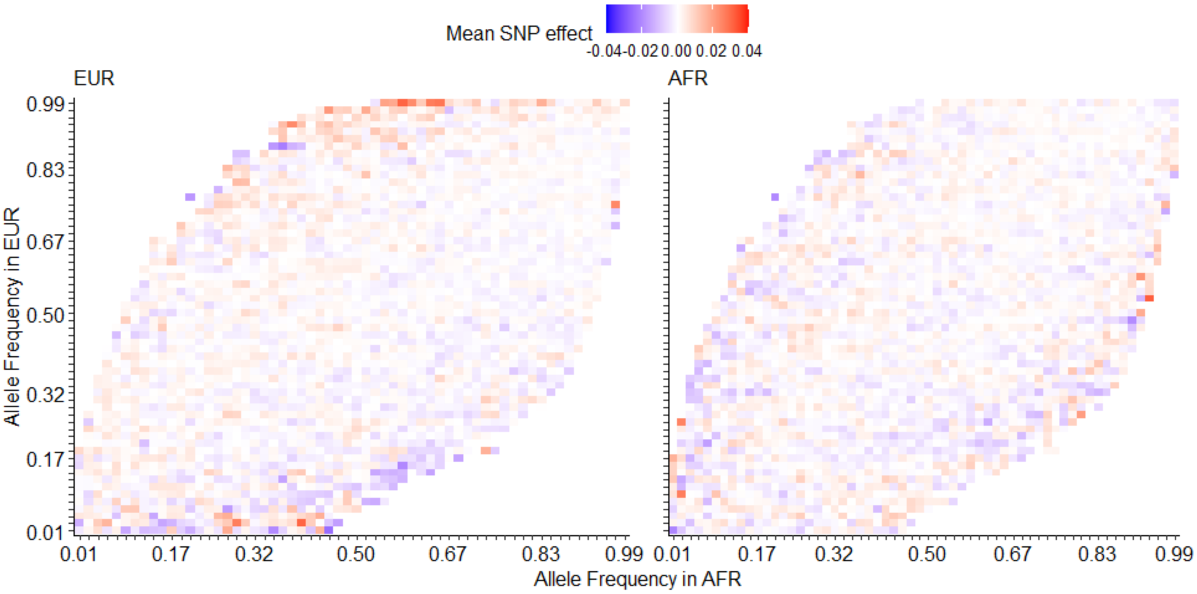


**B. IL-1β**

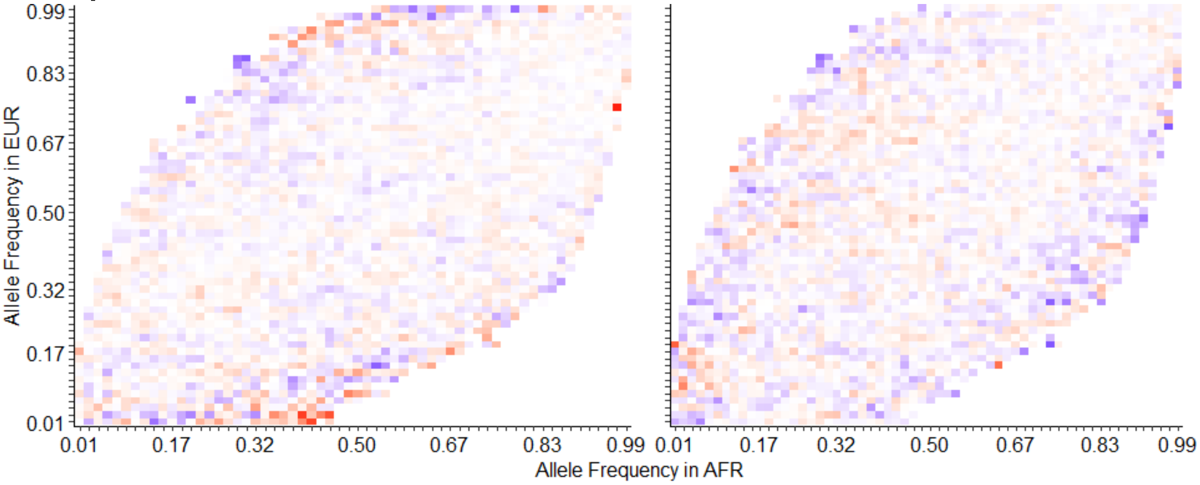


**C. IL-18**

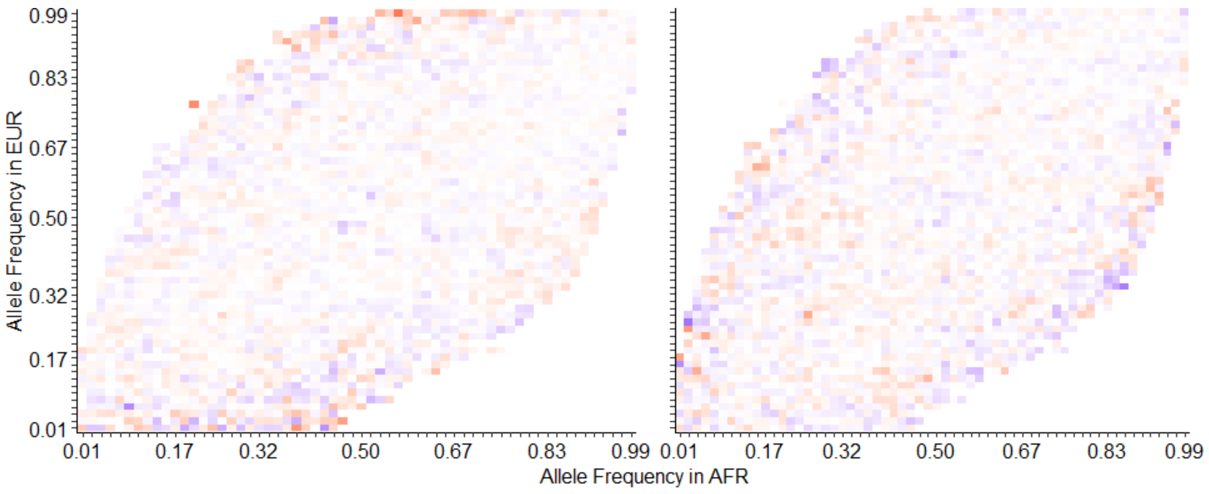

Allele Frequency in EUR
0.99
0.83
0.67
0.50
0.32
0.17
0.01
0.01
0.17
0.32
0.50
0.67
0.83
0.99
Allele Frequency in AFR

**Supplementary Fig. 13. Scatter plot of SNPs from the multivariable Mendelian randomization (MR) model for NLRP3-inflammasome cytokines and atherosclerotic cardiovascular disease (ASCVD).** Each point represents a single-nucleotide polymorphism (SNP) contributing to the inverse-variance weighted MR odds ratios (ORs) for incident ASCVD per genetically predicted increase in plasma IL-6 (**A**), IL-1β (**B**), or IL-18 (**C**) levels, using cytokine-associated genetic variants as instrumental variables. Red triangles indicate individual SNPs associated with increased ASCVD risk while blue triangles indicate SNPs inversely associated with ASCVD risk, with effect size and relevance to vascular disease and/or HIV shown in **Table 2**. Models were adjusted for age, sex, and the first five principal components (PCs). SNPs were selected as strong instruments ($p < 1e-05$ for association with plasma cytokines, minor allele frequency >1%, F-statistic >10) and were linkage disequilibrium (LD)-pruned (250 kb window, $R^2 < 0.5$). Variants with significant individual causal effects are highlighted below: red indicates SNPs increasing ASCVD risk, and blue indicates protective SNPs. The fitted blue regression line represents the overall multivariable MR estimate. Analyses were performed within a case-cohort study stratified by ancestry of 64 ASCVD cases and 283 controls in EUR and 32 ASCVD cases and 294 controls in AFR.

**A.** **IL-6 (EUR)** **IL-6 (AFR)**

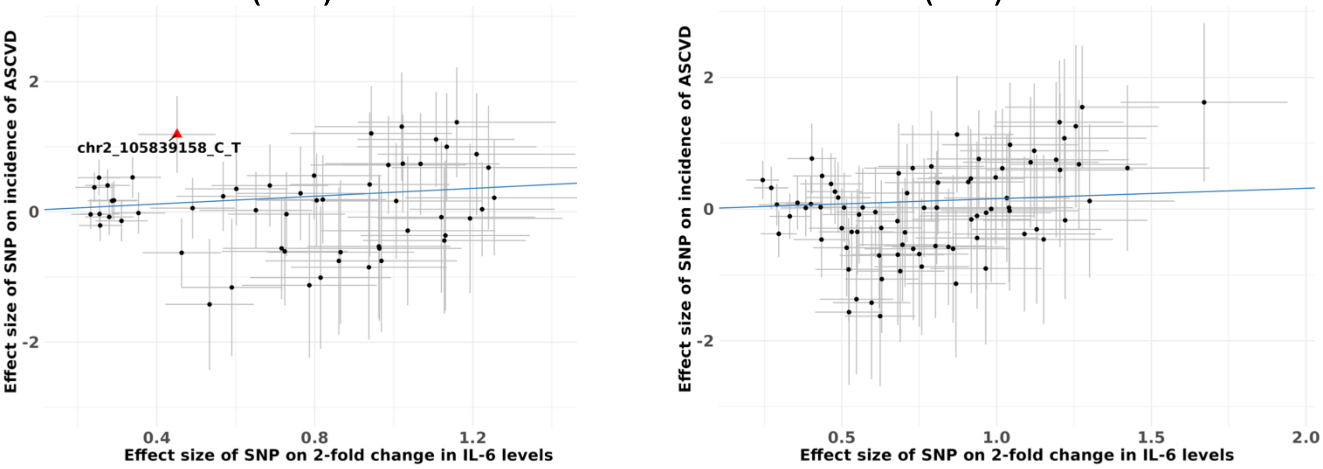


**B.** **IL-1β (EUR)** **IL-1β (AFR)**

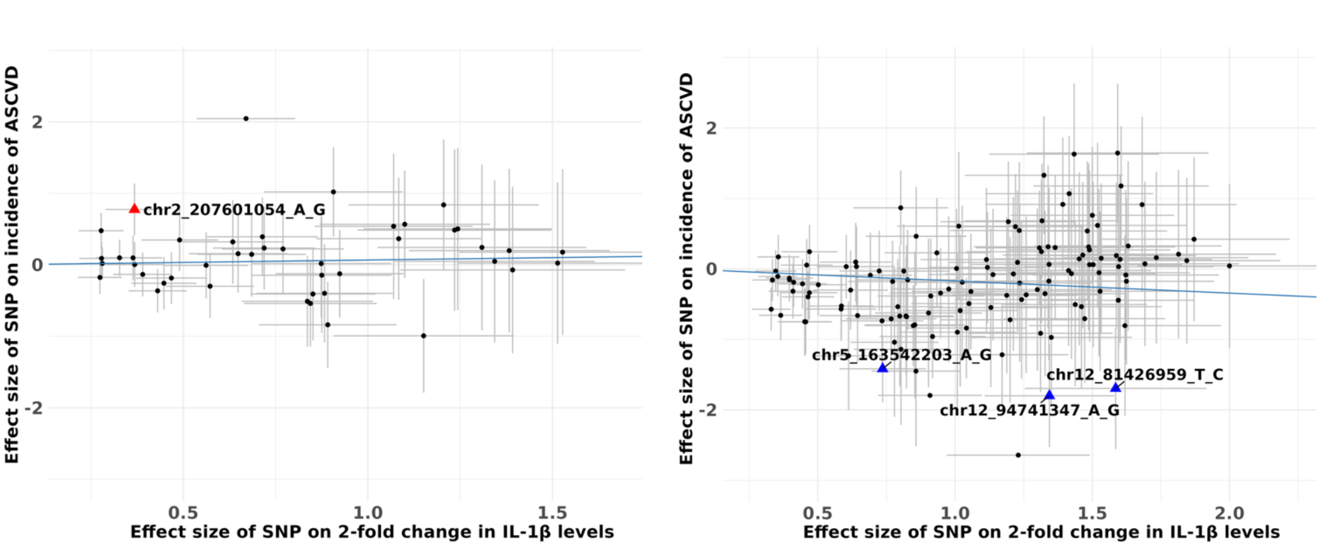

C.

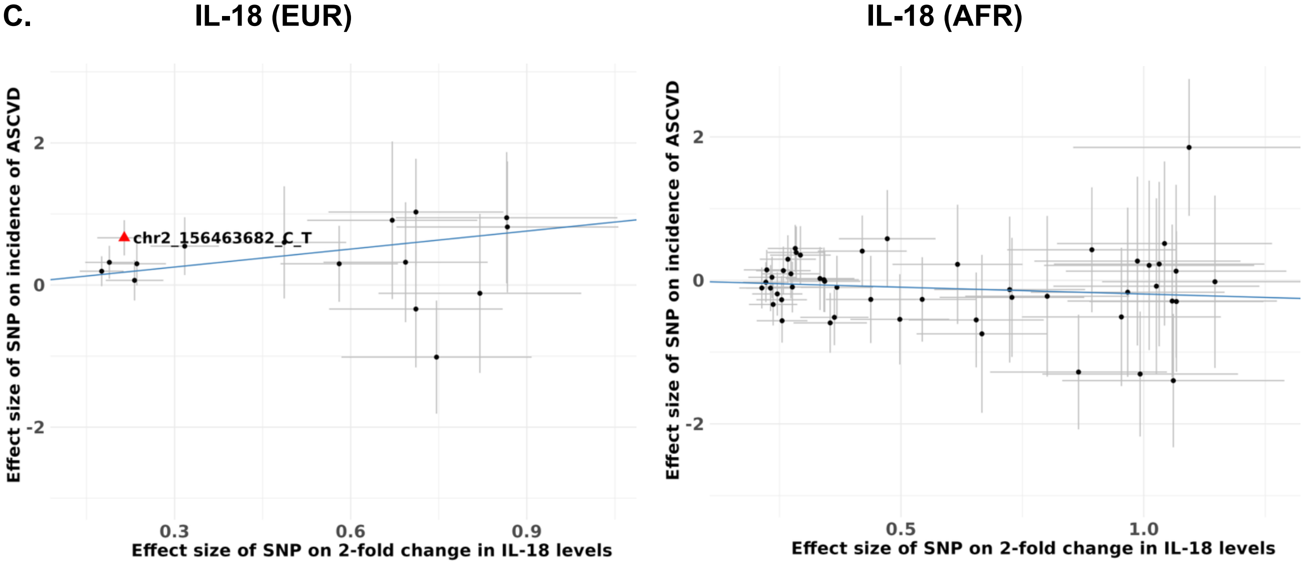
IL-18 (EUR)
chr2_156463682_C_T
Effect size of SNP on incidence of ASCVD
Effect size of SNP on 2-fold change in IL-18 levels
2
0
-2
0.3
0.6
0.9
IL-18 (AFR)
Effect size of SNP on incidence of ASCVD
Effect size of SNP on 2-fold change in IL-18 levels
2
0
-2
0.5
1.0

**Supplementary Fig. 14. Cytokine-associated genomic hits cross-referenced with genome-wide CRISPRa and CRISPRn screens of HIV host dependency pathways in primary human CD4⁺ T cells.** Genes from GWAS, rare variant (RV), and TWAS analyses associated with plasma IL-6, IL-1β, or IL-18 were cross-referenced against gene-level statistics from a published CRISPRa/CRISPRn screen in primary human $CD4^+$ T cells,[89] in which CRISPR-perturbed cells were challenged with GFP-tagged HIV and sorted by FACS into HIV-GFP+ (infected) and HIV-GFP− (uninfected) bins; gene-level log2 fold-change (LFC) and FDR were computed by MAGeCK as the log2 ratio of sgRNA abundance between bins. Bar plots show effect of each candidate gene on HIV infection in CRISPRa (**A**) and CRISPRn (**B**) screens. In both panels, bars are oriented so that green (positive y) = host defense factor (the gene restricts HIV infection) and red (negative y) = host proviral factor (the gene promotes HIV infection). The y-axis shows MAGeCK gene-level log2 fold-change (LFC, log2 ratio of sgRNA abundance between HIV-$GFP^+$ and HIV-$GFP^-$ bins); for CRISPRa, the sign of the displayed LFC was inverted relative to the original MAGeCK output so that bar direction consistently reflects biological function (defense vs. proviral) across both panels. ** FDR < 0.2; * p < 0.05 (computed using the original MAGeCK statistics).

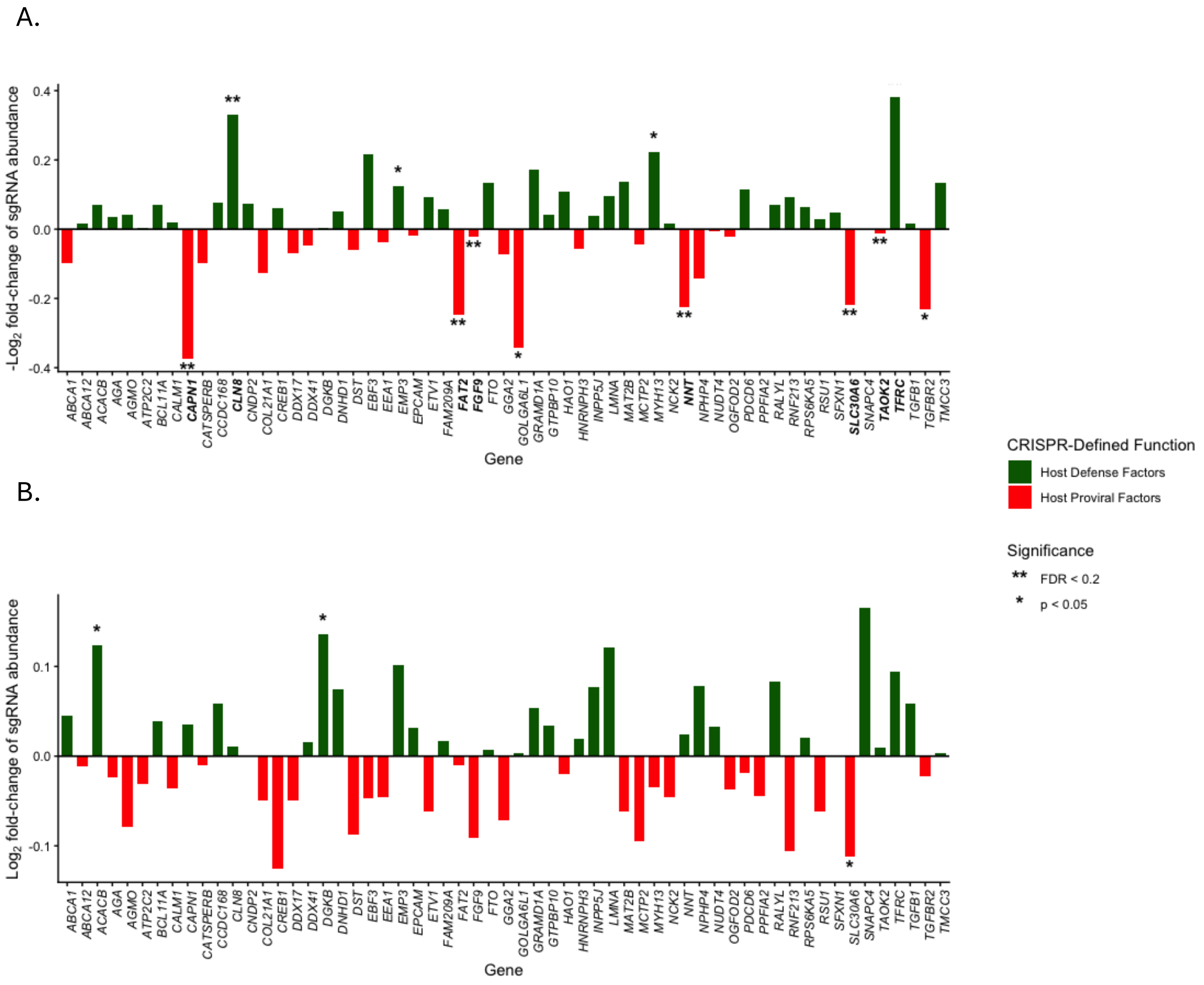

**Supplementary Fig. 15. Validation of IL-6 genetic associations using data from All of Us project.** Publicly available genetic and plasma IL-6 phenotype data from the All of Us Research Program were analyzed using the same analytical pipeline as in our primary study. **(A)** Principal component analysis (PCA) plot of All of Us participants. PC1 and PC2 were used to assign participants to ancestry groups, and individuals mapping to the African (AFR) and European (EUR) reference clusters were selected for downstream GWAS analyses. **(B)** Quantile-quantile (Q-Q) plots of observed versus expected $-\log_{10}$(P-values) from the genome-wide association analyses of plasma IL-6 in the EUR (left) and AFR (right) populations, demonstrating appropriate control of population stratification and minimal genomic inflation. (C, D) Manhattan plots displaying $-\log_{10}$(P-values) from GMMAT-based GWAS of plasma IL-6 across autosomal chromosomes in the EUR **(C)** and AFR **(D)** cohorts. The red dashed line indicates the genome-wide significance threshold (P = 5e-08), and the blue dashed line indicates the suggestive significance threshold (P = 1e-05) Labeled and highlighted points correspond to candidate genes identified at P < 1e-05 in our HIV cohort, shown here at their corresponding positions and significance levels in the All of Us validation cohort for the EUR and AFR ancestry groups.

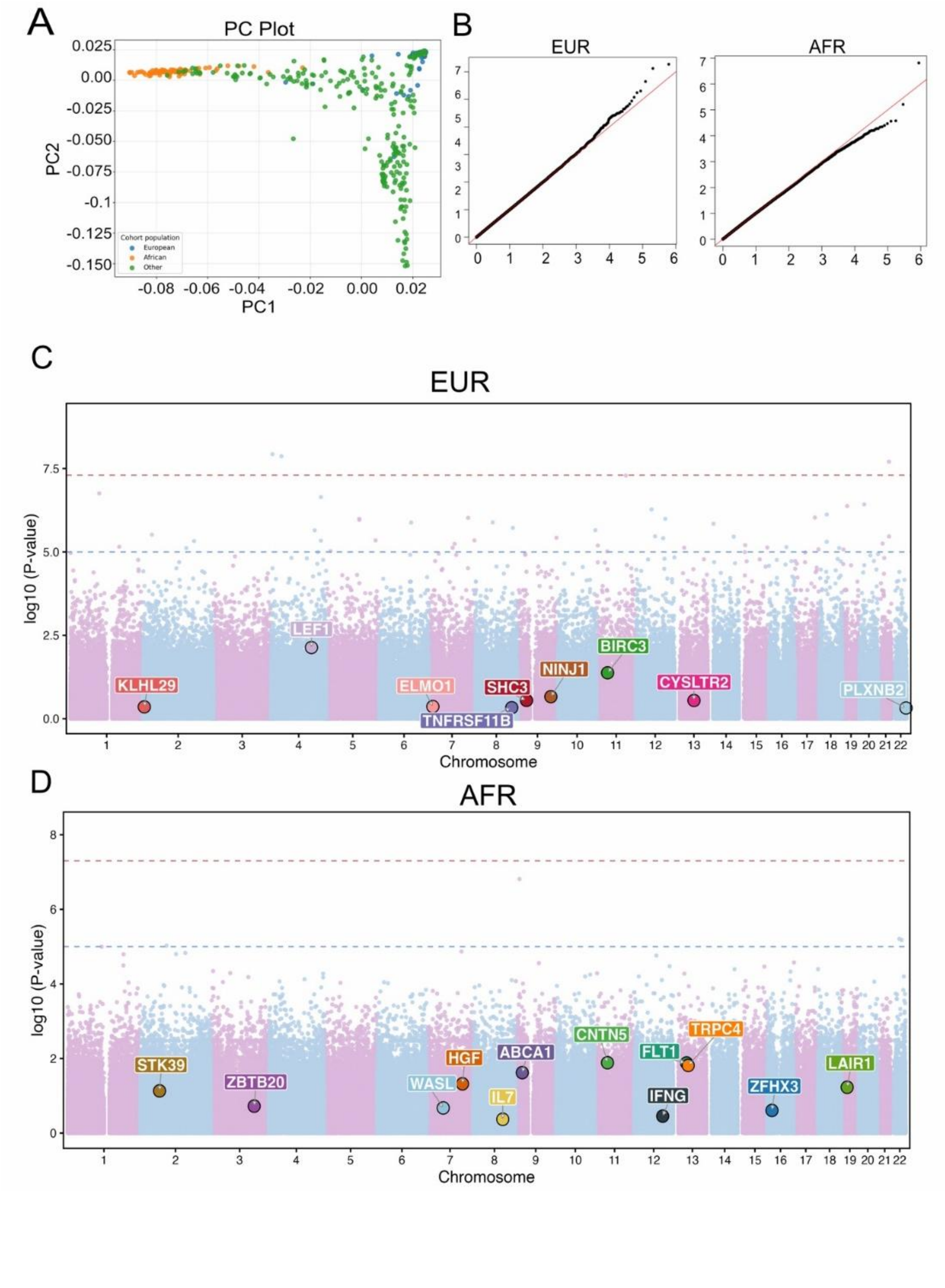
